# Development of models of care coordination for rare conditions: A qualitative study

**DOI:** 10.1101/2021.11.16.21266395

**Authors:** Holly Walton, Amy Simpson, Angus I.G. Ramsay, Amy Hunter, Jennifer Jones, Pei Li Ng, Kerry Leeson-Beevers, Lara Bloom, Joe Kai, Maria Kokocinska, Alastair G Sutcliffe, Stephen Morris, Naomi J. Fulop

**Affiliations:** Department of Applied Health Research, University College London, Gower Street, London WC1E 6BT, UK; Genetic Alliance UK, Creative Works, 7 Blackhorse Lane, London, E17 6DS; Department of Applied Health Research, University College London, 1-19 Torrington Place, London, WC1E 7HB, UK; Alstrom Syndrome UK, 4 St Kitts Close, Torquay, Devon, TQ2 7GD; The Ehlers-Danlos Society and Academic Affiliate Professor of Practice in Patient Engagement and Global Collaboration (Penn State College of Medicine); Centre for Academic Primary Care, NIHR School for Primary Care Research, University of Nottingham, Division of Primary Care, Floors 13-15, Tower Building, University Park, Nottingham, NG7 2RD; Birmingham Women’s and Children’s NHS Foundation Trust; UCL and Great Ormond Street Institute of Child Health, 30 Guilford Street, London, WC1N 1EH; Primary Care Unit, Department of Public Health & Primary Care, University of Cambridge, Cambridge, UK

**Keywords:** Care Coordination, Rare Conditions, Rare Diseases, Qualitative, Health care organisation

## Abstract

**Introduction:** Improving care coordination for people with rare conditions may help to reduce burden on patients and carers and improve the care that patients receive. We recently developed a taxonomy of different ways of coordinating care for rare conditions. It is not yet known which models of care coordination are appropriate in different situations. This study aimed to: i) explore what types of care coordination may be appropriate in different situations, and ii) use these findings to develop hypothetical models of care coordination for rare conditions.

**Methods:** To explore appropriateness of different types of care coordination, we conducted interviews (n=30), four focus groups (n=22) and two workshops (n=27) with patients, carers, healthcare professionals, commissioners, and charity representatives. Participants were asked about preferences, benefits and challenges, and the factors influencing coordination. Thematic analysis was used to develop hypothetical models of care coordination. Models were refined following feedback from workshop participants.

**Results:** Stakeholders prefer models of care that: are nationally centralised or a hybrid of national and local care, involve professionals collaborating to deliver care, have clear roles and responsibilities outlined (including administrative, coordinator, clinical and charity roles), provide access to records and offer flexible appointments (in terms of timing and mode). Many factors influenced coordination, including those relating to the patient (e.g., condition complexity, patient’s location and ability to coordinate their own care), the healthcare professional (e.g., knowledge and time), the healthcare environment (e.g., resources) and societal factors (e.g., availability of funding). We developed and refined ten illustrative hypothetical models of care coordination for rare conditions.

**Conclusion:** Findings underline that different models of care coordination may be appropriate in different situations. It is possible to develop models of care coordination which are tailored to the individual in context. Findings may be used to facilitate planning around which models of care coordination may be appropriate in different services or circumstances. Findings may also be used by key stakeholders (e.g. patient organisations, clinicians and service planners) as a decision-making tool.

## Introduction

Patients and family members are increasingly expected to be involved in the day-to-day management and organisation of their care, due to increased demands on healthcare services and a shift in accountability of healthcare [1]. This is particularly true for patients and families living with rare conditions. Rare conditions (including ultra-rare and undiagnosed conditions) are defined as those which affect up to five in every 10,000 people [2,3], affect many different body systems [3,4], and require care from a range of professionals and sectors. Previous research has found that care for people with rare conditions is often not coordinated, resulting in them attending multiple appointments, on different days, with different professionals in different locations [2,5,6]. Additionally, patients with rare conditions often do not have a designated care coordinator [6–9], and thus the role of coordinating care frequently falls to patients and carers [7,8]. Within this role, patients and carers often undertake tasks such as chasing and organising appointments, chasing test results and passing information between different healthcare professionals [8].

Previous research has demonstrated the potential benefits of improving care coordination for people with rare conditions. For example, research highlights the negative physical, psychological, social and financial implications that a lack of coordination can have for patients and families living with chronic and rare conditions [7,10,11]. Additionally, it is widely thought that improving care coordination across a range of common and rare conditions may lead to improved outcomes for patients and healthcare systems [2,5,6,12]. This is reflected in recent UK policy initiatives to improve care coordination for patients with rare conditions [2,5,13,14].

A scoping review of reviews of common and rare chronic conditions defined care coordination for rare conditions. Coordination should be family-centred, evidence-based, and equitable and should involve all of those involved in a person’s care working together to achieve the same goals and outcomes across a person’s whole life, and across all sectors [8].

Findings from previous research relating to chronic conditions [1,8,16], together with the vast number of rare conditions, differences in availability of services (e.g. highly specialised services have been commissioned for some rare conditions, but specialist centres are not available for all conditions or patients [6,9]), and diversity of experiences existing within the rare disease community indicate that there are likely to be a range of factors that may influence coordination. Factors influencing coordination for rare conditions have not yet been fully explored.

We have recently developed a taxonomy of care coordination for rare conditions. The taxonomy outlines six domains of care coordination, each with a range of options for coordinating care: i) ways of organising care (national, hybrid and local), ii) ways of organising individuals involved in a person’s care (collaboration between many/all, some or no professionals), iii) responsibilities (administrative roles, formal roles: coordinator, clinical lead, GP, and supportive roles: charities, patients/carers), iv) how often appointments and care coordination take place (regular, on demand and hybrid), v) access to records (full or restricted for patients and healthcare professionals) and vi) mode of communication (digital, face-to-face, phone) (see Appendix 1 for summary; or [15] for further details).

Whilst previous research has outlined the different domains and options for coordinating care for rare conditions [15], we do not yet know which options and models of care coordination stakeholders prefer and which models may be appropriate in different situations. This study aimed to: 1) explore what types of care coordination may be appropriate in different situations, and 2) use these findings to develop hypothetical models of care coordination for rare conditions.

The manuscript outlines the methods and findings in two stages: (i) exploring what types of care coordination may be appropriate in different situations, and (ii) developing hypothetical models of care coordination.

## Methods

### Design

This study is part of a wider mixed-methods research project which explored coordination of care for people with rare conditions [11].

#### Aim 1. Exploring what types of care coordination may be appropriate in different situations

This manuscript builds on previous research which outlined the development of a taxonomy of care coordination for rare conditions (see Appendix 1 for summary, or [15] for details). As part of this study, interviews, focus groups and workshops were conducted to explore what types of coordination may be appropriate in different situations.

To explore what types of care coordination may be appropriate in different situations, we explored the following aspects: stakeholder preferences for different types of care coordination, benefits and challenges of using different types of care coordination, factors influencing the use of different types of care coordination, and barriers and facilitators to coordinating care more generally (see Figure 1).

**Figure 1.**
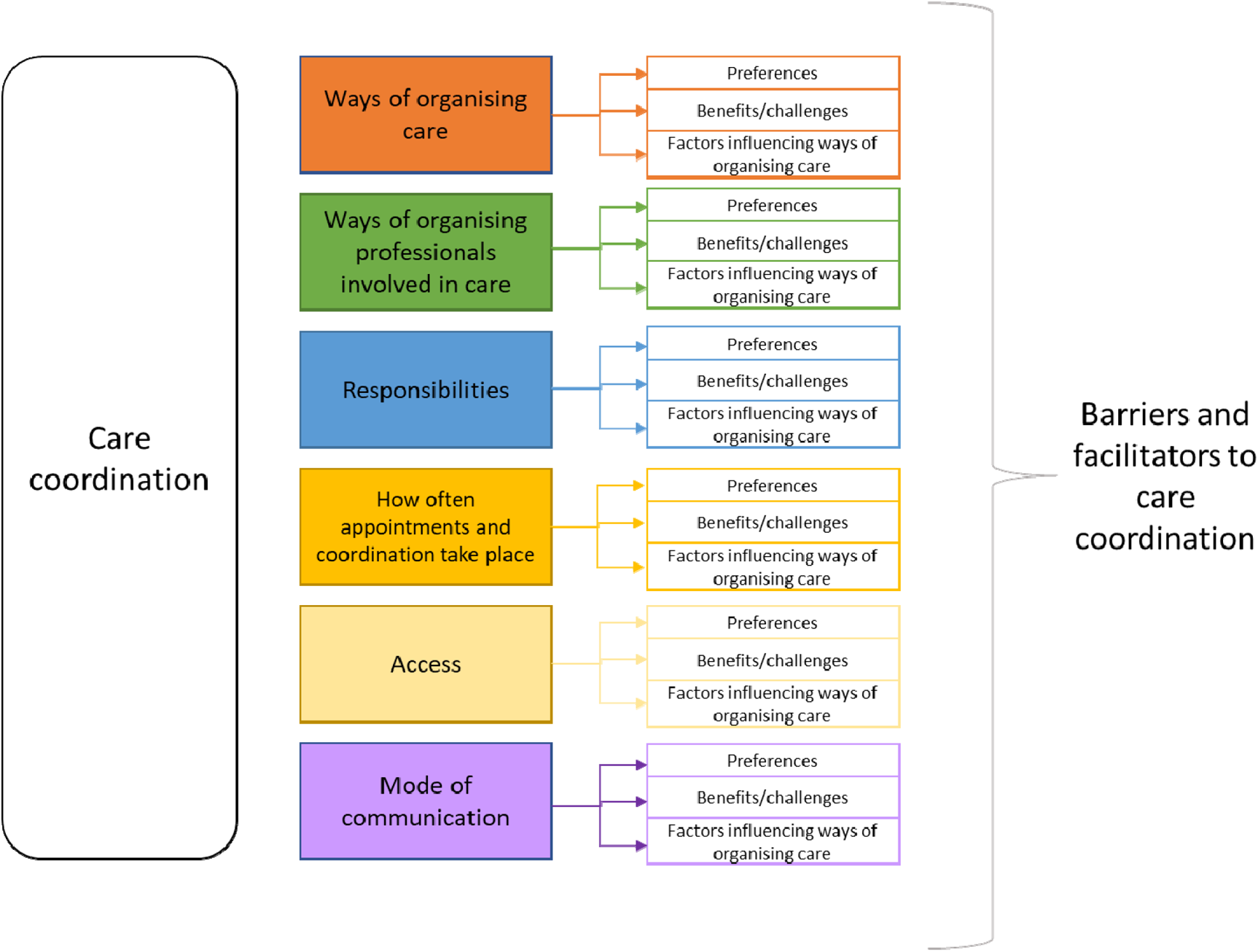
A summary of the topics explored in this study

The methods for conducting these interviews, focus groups and workshops are reported in detail in Appendix 2 (or see [15]).

##### Sample

This study included 79 participants (patients, carers, healthcare professionals, commissioners and charity representatives). This included 30 interview participants (healthcare professionals/charity representatives/commissioners), 22 focus group participants (patient/carers) and 27 workshop participants (12 patients/carers, 15 professionals) (see Appendix 2 for further details).

##### Measures

We developed topic guides for interviews and focus groups (see Appendix 3, or [15]). This manuscript will draw on the analysis of data relating to experience of different types of care coordination (including preferences, benefits and challenges and factors influencing coordination) and barriers and facilitators to coordinating care generally (see Figure 1). During the workshops we also asked participants for their feedback on appropriateness of the options presented in light of COVID-19 (see Appendix 3, or [15]).

##### Procedure

One researcher (HW) conducted 30 interviews (ranging from 44 minutes to 74 minutes) with healthcare professionals, commissioners, and charity representatives by telephone (n=27) or in person (n=3). Two researchers (HW/AS) conducted four focus groups (ranging from 149-154 minutes) with patients and carers either in person (n=2; in two UK locations) or virtually using Skype for business (n=2). We recorded interviews and focus groups, and transcripts were professionally transcribed. We checked transcripts for accuracy and fully anonymised names, places and specific conditions.

Following initial analysis of the interviews and focus groups, we held two workshops to validate and build on interim findings. Workshops were held online and were recorded. Notes were checked, and summarised. Notes were sent to a graphic facilitator (New Possibilities) to create a graphical representation of the findings. Data were stored in UCL’s Data Safe Haven and coded using NVivo 12.

The procedure for collecting interview, focus group and workshop data used in this study are described in depth in Appendix 2 (and [15])

##### Analysis

Given the large amount of data in this study, analysis was conducted in two stages: 1) development of themes and sub-themes for the data on aspects of coordination (to develop initial taxonomy options) (described in [15]), 2) development of themes and sub-themes relating to appropriateness of different care coordination models in different situations (described in this manuscript; see Figure 1).

Inductive coding was used to develop an initial coding frame [18]. Six transcripts were coded inductively by two researchers (HW/AS) and a coding framework was then developed and agreed. The coding frame included codes relating to different options of coordinating care (see [15]) and also codes relating to participant preferences for different types of care coordination, benefits and challenges of different types of care coordination, factors influencing different types of care coordination and barriers and facilitators to coordinating care more generally. This coding frame was then used to deductively code all interview and focus group transcripts (HW) [19]. A second researcher (AS) coded six interviews and one focus group transcript (20% of data) and coding was discussed and agreed.

We then conducted an analysis of findings relating to preferences, benefits/challenges, factors influencing coordination and barriers and facilitators for each of the six taxonomy domains (see Appendix 1) using the iterative categorisation process [19]. For example, for factors influencing coordination, we developed themes and sub-themes, including patient factors (e.g., diagnosis, age, condition, individual patient needs and preferences, consent, ability to travel), healthcare professional factors (e.g., knowledge and understanding, skills and capability, attitudes, opportunity) healthcare environment factors (e.g., resources, environment, attitudes), and societal factors (e.g. resources/funding). We coded data in relation to preferences, benefits/challenges, factors influencing coordination for different coordination options within these categories.

To supplement our analysis, we coded and grouped workshop notes into themes surrounding experiences of different models of coordination, benefits and challenges of the models of coordination, factors influencing coordination, missing aspects, and impact of COViD-19.

#### Aim 2. Development of hypothetical models of care coordination for rare conditions

Once we had identified stakeholder preferences, benefits and challenges of different models, factors influencing coordination of different models and barriers and facilitators, we used these findings to develop hypothetical models of care coordination which outline options for coordinating care in different situations. These were based on different combinations of domains and options described in our taxonomy [15].

We developed the hypothetical models of care coordination in three stages: i) development of the CONCORD flow chart (see Appendix 4), ii) development of illustrative models, and iii) refinement of illustrative models. The procedure for each of these three stages is described in Figure 2.

**Figure 2.**
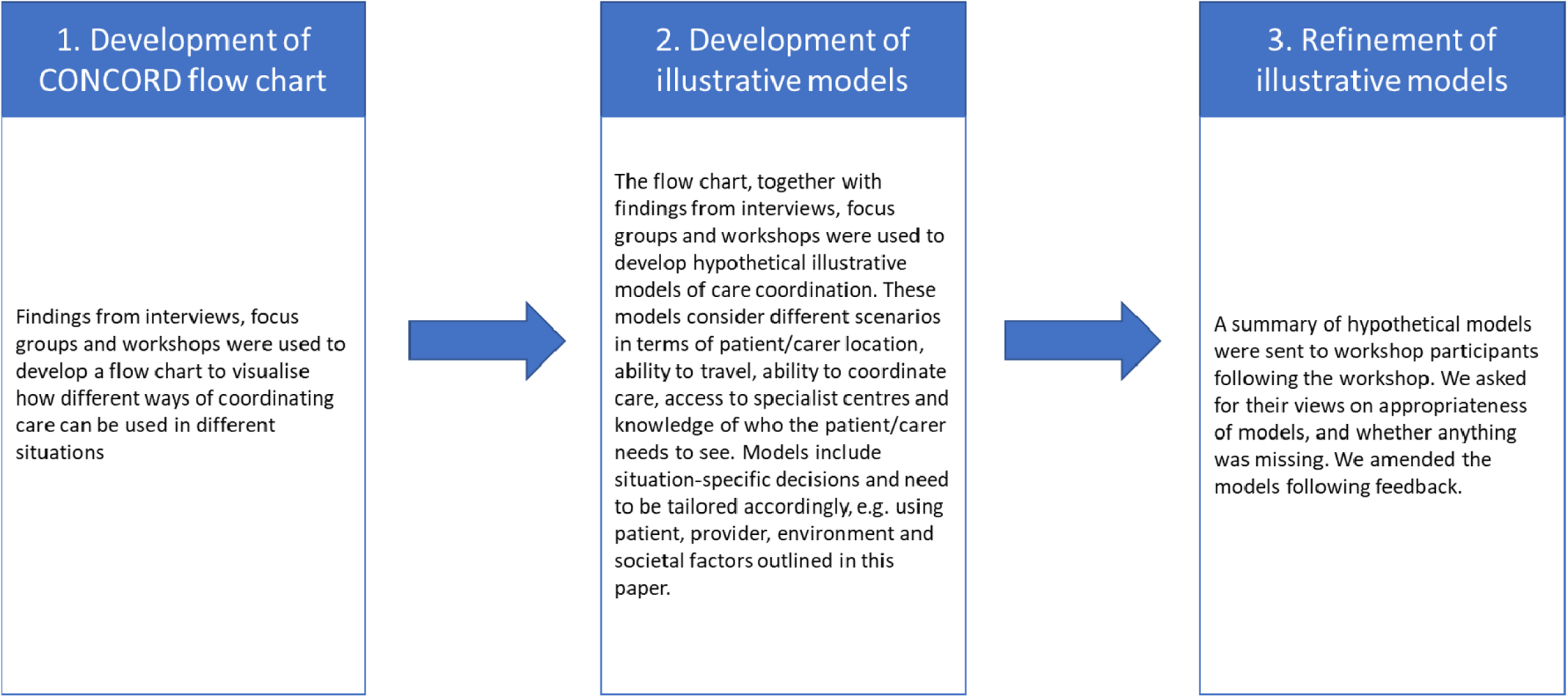
Summary of the process used to develop hypothetical models of care coordination

##### i) Development of the CONCORD flow chart

The CONCORD flow chart was developed using the taxonomy (which outlines examples of different ways of coordinating care in practice) and qualitative findings on care coordination (preferences, benefits/challenges, factors influencing coordination and barriers/facilitators) from 30 interviews with healthcare professionals, commissioners, charity representatives and four focus groups with patients and carers). The CONCORD flow chart is a visual representation of the findings presented in this manuscript and in Walton et al [15]. One researcher (HW) developed the CONCORD flow chart to visualise how the different ways of coordinating care can be used in certain situations.

The CONCORD flow chart includes the six domains from the CONCORD taxonomy. We included all six domains as they were all found to be important when coordinating care. Within the flow chart, a series of questions are asked to help users to think about which option of coordination may best suit patient, family and service circumstances. The flow chart has decision boxes (boxes that are fully shaded). Within each decision box, there are multiple options that may be suitable (e.g., the type of technology, mode of communication, or who coordinates care). The flow chart is not designed to account for all possible situations, but instead aims to support discussion and thinking around which models may suit different situations. In addition to the flow chart (see Appendix 4), we have also designed a cover note to help users to understand how the flow chart can be used (see Appendix 5).

##### ii) Development of illustrative models

Using the CONCORD flow chart and the taxonomy [15], we developed some hypothetical illustrative models of care coordination. These were designed to illustrate the use of the taxonomy and the CONCORD flow chart. We developed hypothetical models instead of actual care coordination models as the findings indicated that there were many different ways care could be coordinated, and that we may not be able to fully represent all situations, domains and options of care coordination if using real life examples. However, many real-life examples of different ways of coordinating care are shown in Walton et al [15].

To develop the illustrative models, we considered different scenarios in terms of: i) where the patient and parent/carer lives in relation to a specialist centre, ii) whether the patient and parent/carer can or wants to travel to a specialist centre; iii) whether the patient and parent/carer has the ability (and wants to coordinate their own care), v) whether the patient and parent/carer has access to a specialist centre, vi)) whether it is clear who the patient needs to see for management of the condition.

Eight models were initially developed (including models for conditions that have access to specialist centres, and models for conditions which do not have access to specialist centres.

When developing the models, we also highlighted how additional situation-specific decisions (based on the factors influencing coordination) would need to be considered within each model (e.g. the level of coordinator support available and needed; who the coordinator is and who the clinical lead is; who should be involved in MDT meetings; the extent to which different modes are used for information sharing, communication, care delivery and coordination; the extent to which information is shared; the extent to which providers have access to records; how often care coordination and care appointments are needed; and, transition needs).

##### iii) Refinement of illustrative models

To refine the models, we sent a handout summarising the hypothetical models to CONCORD workshop participants (patients, carers, healthcare professionals, commissioners and charity representatives) who consented to provide feedback. We asked them for their views on whether the models seemed appropriate based on their experiences, and why, and whether we had missed any obvious models of coordination.

We received written feedback from eight workshop participants and members of the CONCORD research team. To address the feedback and refine the models, we grouped the feedback into two categories: ‘feedback on the models’ and ‘suggested improvements’.

Findings indicated positive feedback about the hypothetical models, but highlighted that they may not currently be seen in practice and/or be feasible, but that models should be aspired to in future.

Feedback informed a range of amendments. These included: adding transition into all models; broader use of digital and remote technologies; formal shared care models; clarifying that who is involved in outreach clinics varies, emergency healthcare planning; signposting patients with undiagnosed/ultra-rare conditions to patient support groups; arranging appointment frequency based on need and explaining the role of care coordinators. Further models of coordination for those without access to a specialist centre was also included.

Amendments resulted in ten hypothetical models of care coordination.

## Results

### Aim 1. Exploring what types of care coordination may be appropriate in different situations

In this manuscript we present findings relating to the appropriateness of different types of care coordination in different scenarios, in relation to the six taxonomy domains (see Appendix 1, or [15] for further details): 1) ways of organising care, 2) ways of organising the team, 3) responsibility for coordination, 4) how often appointments and coordination take place, 5) access, and 6) mode of information sharing, consultation and communication.

Table 1 outlines example quotes relating to preferences, benefits and challenges and factors influencing coordination for different types of care coordination.

**Table 1.** Example quotes for each of the domains and contextual factors

| Domain (from Walton et al [15]) | Contextual factor | Example quote |
| --- | --- | --- |
| 1. Ways of organising care | Preferences | *"Hmm... well obviously ideally close to home but I think the majority of our patients, if they feel they're going to be get a good service and a specialist service, they are willing to travel to a specialist centre." (Interviewee, healthcare professional)* |
|  | Benefits/challenges | *"...they've become specialists in that particular condition, and they know what they're talking about and they know...you know, the advice that they give you, is the correct advice for that condition. So I think in some ways, going up there is important because you know that you're receiving the right kind of treatment and advice." (Focus group participant, Patient)*<br>*"[Rare condition 1] was lucky because it got in there at the beginning but you can't have a separate service for every single one" (Interviewee, healthcare professional)*<br>*"I think it's about good communication, it keeps communication links open if you have a named therapist in each locality. I think it's about feeling supported. I think it's about shared learning. So if you're working closely with therapists in the more hub roles, we're reliant on them feeding information back to us on how the patients are doing and they're reliant on us a) making them the referrals, and b) advising them on treatment." (Interviewee, healthcare professional)* |
|  | Factors influencing coordination | *"so it's more, you know, a one-stop shop to try and get everything done in one go. But actually that model often only works well if you've got a very discreet phenotype, you know, a discreet medical condition that you're looking at rather than it being something that is going to work for everybody because if you don't know what's going on it's difficult to know who that individual needs to see when they come in for assessment" (Interviewee, healthcare professional)*<br>*"You see, for me, I would like to have someone who's consultant-level but not necessarily specialised, but is at least willing to learn, that I can go to, because travelling is something that's such a big issue for me [...] So for me, it would be nice to be able to go to someone locally who could co-ordinate and just be my go-to person and...or that also my local hospital can bring in when I'm admitted, so that I get some continuity of care, because I go through the same thing every time, to the point now where I actually am scared of hospitals, scared of doctors" (Focus group participant, Patient)* |
| 2. Ways of organising professionals involved in a patient's care | Preferences | *"we do support this idea of multidisciplinary team clinics, and then that those MDTs develop good lines of communication with GPs and other providers. That seems to work the best, and we think that there's some evidence that patients do better when they're under the care of those sorts of clinics." (Interviewee, charity representative and healthcare professional)*<br>*"And, you know, I feel like if he had a clinic appointment where he could go to and everybody was there, so they had speech and language, they had physio, they had OT, everyone, you know, was there in the clinic, they could see him and then arrange further appointments. I feel that would just be so much more beneficial for us because, you know, having a young family is hard enough, having a child with, you know, a disability is hard enough, but put that altogether with the frustrations that come from a lack of coordination from your healthcare it then adds more stress because you do end up feeling like he's falling through the net and it does, he gets missed a lot and you end up phoning and that for me is a real frustration." (Focus group participant, Parent/carer)*<br>*"There should be early introduction to the concept of transition and then plan it with both paediatric and adult services." (Interviewee – commissioner)* |
|  | Benefits/challenges | *"... because we're all there on hand in clinic, we can then, you know – the patient can be directed to the appropriate, sort of, allied healthcare professional to address that particular need. So, it works, you know, it works – or if one of us weren't there or if we're busy with another patient, you know, at the MDT meeting we can make sure that a follow up call for all appointments happen, depending on what the emerging needs are, really. (Interviewee – healthcare professional)* |
|  | Factors influencing coordination | *"I guess, by the nature of it, it is that complexity that there are so many people involved, and, yeah, who is making that decision, who makes the ultimate decision and who is the right person to coordinate that, I think, and time and money is always a factor, you know, having somebody to find to have that role to coordinate and have the time to do that. You know, it's fine when you've got a few patients (inaudible 0:47:59), but we just don't have that built into our timetables to do that extra liaison that you need sometimes." (Interviewee – healthcare professional)* |
| 3. Responsibilities | Preferences | *"I agree. I don't think it's difficult. I think you can have a... I think you need a named consultant as the overall co-ordinator... [...] then maybe the person you have face-to-face with, the person who is a co-ordinator or... Often I do think the senior nurse is really good." (Focus group participant, patient)*<br>*"You know, I appreciate that GPs are incredibly busy and they have a whole host of thousands of patients to consider under their care. However, I do think they have to take some kind of responsibility to some degree to know that every patient under their care could be struggling with all different kinds of things, and I think if it's a particular one that we could help with that is a noted rare disease, then I think it's important that we liaise with them and make sure that they're aware of all the different services that are available locally, and also that they can offer to the patient, because they are the referrer or care, as well." (Interviewee – healthcare professional)*<br>*"I think sometimes in an ideal world they, you know, they'd like you to be sitting at the end of the phone available for you – for them there and then, and sometimes that – you know, sometimes they get lucky and you are at your desk and you can take the call. I think our email system works well in that – I mean, we're not able to, sort of, answer the queries or address them necessarily straight away, and so we do make it clear that it's not for urgent urgent things, but it's a way of things coming in centralised and then the queries can get triaged out to the, you know, the most appropriate member of the team." (Interviewee – healthcare professional)*<br>*"Oh, I think they're immensely important. Working with three who link to all of our clinics they are fantastic because they do help with coordinating and they are often people who are overlooked in terms of being the – it could be that they could have this role of supporting coordinators. I don't think it should fall to them completely because unless their job role is to do that they wouldn't have capacity to do that, but they are certainly part of that triangle of education, health, and social care because sometimes you get a Family Support Worker who will know quite a lot about the educational input but you might not have a nominated person from education to speak on their behalf so the Family Support Worker can do that and then linked to health and social care. So, I think their role is brilliant." (Interviewee – healthcare professional, speaking about charity involvement)* |
|  | Benefits/challenges | *"Well, I think where they've got them then it makes a huge difference [...] where they have perhaps got one of the hospice neuro nurses or they've got a community matron, you know, they're very happy that their care is really well managed, they're happy that they've got somebody that they can speak to who knows them, who's got that continuity, and who, you know, makes sure that they get to know the other people who can be helpful to them. So, it saves them having all these random contacts from people not necessarily at the most useful time. That person ensures that they get the right input at the right time, and also they're confident that that information is being liaised between people and professionals, and that's quite difficult sometimes if you haven't got a cohesive team and you haven't got somebody taking that central role. So, I think, you know, when you talk to our folks who has got that, they're all the, sort of, positives that they mention." (Interviewee, voluntary sector healthcare professional)* |
|  | Factors influencing coordination | *"I am pretty certain families would say, "We want others to take... we want to be part of it," and clearly they couldn't not be, and they now expect to* |
|  |  | *be part of the co-ordination of their child's care, as do adults of course, now, taking much more... But they don't want to be...I think there's a range of abilities to be in the driving seat, so there are some who don't know what...some are...So many people now are on top of it, you know, they are on top of their own information and will drive the process, and they will go to their...but they still expect their...particularly their surgeons, to drive what's happening with them, and their nurses in those specialist units. I think there's a whole hidden...a whole range of hidden patients, if you like, that we don't know about, and those who are not as articulate and as savvy about their condition, who probably need a lot more handholding, and so I think to make a generalisation of, "What do you think people want from it?" I think it depends on the circumstance of the individual." (Interviewee – charity representative)* |
| 4. How often care appointments and coordination take place | Preferences | *"No, I think that regular appointments every six months are fine as long as you're able to contact somebody in between if there is, you know... If you need to go to A&E, as long as you have that one person that you can contact all the time, they can either send you to a scan, so by the time you go for your six months, you know what the problem is and then you can have something done and move on. Emergency ones are different. I think sometimes you can't really put a time on anything like that because, say, if you have a seizure, or something like that, or a fall, or anything, you can't really put a time on it, you need to be seen straightaway." (Focus group participant, patient)* |
|  | Benefits/challenges | *"which when we were in [Country 2] under the SPZ, that was every three months, which was, for that degree of disability of my daughter, was perfect. So, I think that 12-week, sort of, window is enough to, if something really has changed, if something needs to be caught, it really- that's just- the perfect for us would be that, for those, kind of... then there's, obviously, the reactive who knows what happens in the preceding week, but in terms of that oversight, that, sort of, three months for me would be the perfect figure". (Focus group participant, parent/carer)*<br>*"I find sometimes if you have yearly or six-monthly appointments time and time again, they can be a bit fruitless, you don't really get anything from the appointment, because you're not in there for a specific purpose or reason, they're just going, "Okay, you're fine, see you next year." It might be unnecessary to have that all the time." (Focus group participant, patient)* |
|  | Factors influencing coordination | *"I guess it depends on the condition and how much things are changing, and whether it is a life limiting condition, because if it is a life limiting condition there is probably more things that are changing more rapidly. So, I think it has to be condition specific, so I guess you would be guided by what the experts think is appropriate" (Interviewee – charity representative and healthcare professional)* |
| 5. Access to records | Preferences / benefits and challenges | *"I mean, I personally wouldn't mind it shared with anyone. I'd rather the more people."*<br>*[...]*<br>*I'm the same. I mean, with my daughter, I was, like, "Yay. If you want to look at this and you want to use this to help her, [...] if this can make you more informed, if this can connect you to my daughter, please do ahead and do it," you know." (Focus group participants, patient and parent/carer)*<br>*"Yeah, I think there should be a button where we, if necessary, can delegate the authority for people to read it, say if you've got a problem that's going on a long time, yes, I delegate it, but at the same time, you can also press that button and take that delegated authority back." (Focus group participant, patient)* |
|  | Benefits/challenges | *"It would be lovely if when a child came into our hospital, they came with, well I'd say a little book but we are going paperless, aren't we? But that they would come with some sort of package where you knew their GP, you know their local physio, you knew their speech and language therapist, that it was all related, all in one place. And perhaps if they didn't have local physio services, depending where they lived, what would be their local* |
|  |  | team. Because we spend so much time trying to find that out, that if it was all like through a GP surgery, or just at the very local level, just all that information, if the child came with that, that is what to me co-ordinated care is, making sure that you know everyone that is involved and that you have got open access to those people.” (Interviewee, healthcare professional)<br>“Because I want to know who’s reading my, you know, someone did say at one time, “Oh, the psychiatric team are looking at your notes,” I haven’t given them permission to do that. [...] You know, why are they looking at my notes and for what reason?” (Focus group participant, patient) |
|  | Factors influencing coordination | “And I think the way going forward will be that we’ll all move more electronic and there will be some form of, I don’t know, NHS Cloud that people can log into or something, but there will be something in the digital technology that we’ll all move to. At the moment it is still paper-based and very much letters.” (Interviewee, healthcare professional) |
| 6. Mode of information sharing, consultation and communication | Preferences | “I think that in a totally ideal world – see, it’s pie in the sky, but in a totally ideal world, if all of the NHS had electronic patient records that were all on the same system and could be shared automatically between units then, you know, we’d be able to see things more nationally.” (Interviewee, charity representative and healthcare professional) |
|  | Benefits/challenges | “Having that digital ability to share information, as well, I think would be really invaluable, and making sure that all medical professions are sharing copies of information, letters, to each other so that they all know what’s going on. I think the one thing that I haven’t mentioned yet would be a national portal.” (Interviewee, charity representative)<br>“So yeah, I mean, I guess if, I guess potential models going forward, obviously it’s very much disease dependent, but if you’ve got a patient with a complicated rare disease, where there’s a few national specialist centres, I could envisage a bit where you would have a clinic appointment annually at that specialist centre, so you’ve got those face-to-face, and then a remote appointment at some other interval over the year dependent upon the disease combinations, and that may be in combination of possibly, like, a telehealth appointment with one other member of the healthcare team who would be able to action things locally. [...] And the patient wouldn’t necessarily need to be physically there.” (Interviewee, healthcare professional) |
|  | Factors influencing coordination | “I think it needs to be face-to-face, particularly with, you know, some of these conditions where the person themselves isn’t going to be able to communicate even, you know, by Skype or email or telephone. [...] and I think you need to see what’s happening in that person’s environment, see the pressures that everybody’s under, and actually see the person for yourself to work out where things are at. So, I do think there needs to be capacity for face-to-face. It doesn’t need to be all face-to-face, but you need to be able to have a regular touch base in their own space of what’s happening for them, what are their priorities, what are their challenges” (Interviewee, voluntary sector healthcare professional) |

A summary of findings relating to preferences for different types of coordination, benefits and challenges relating to different types of coordination and factors influencing different types of coordination are shown in Table 2.

**Table 2.**
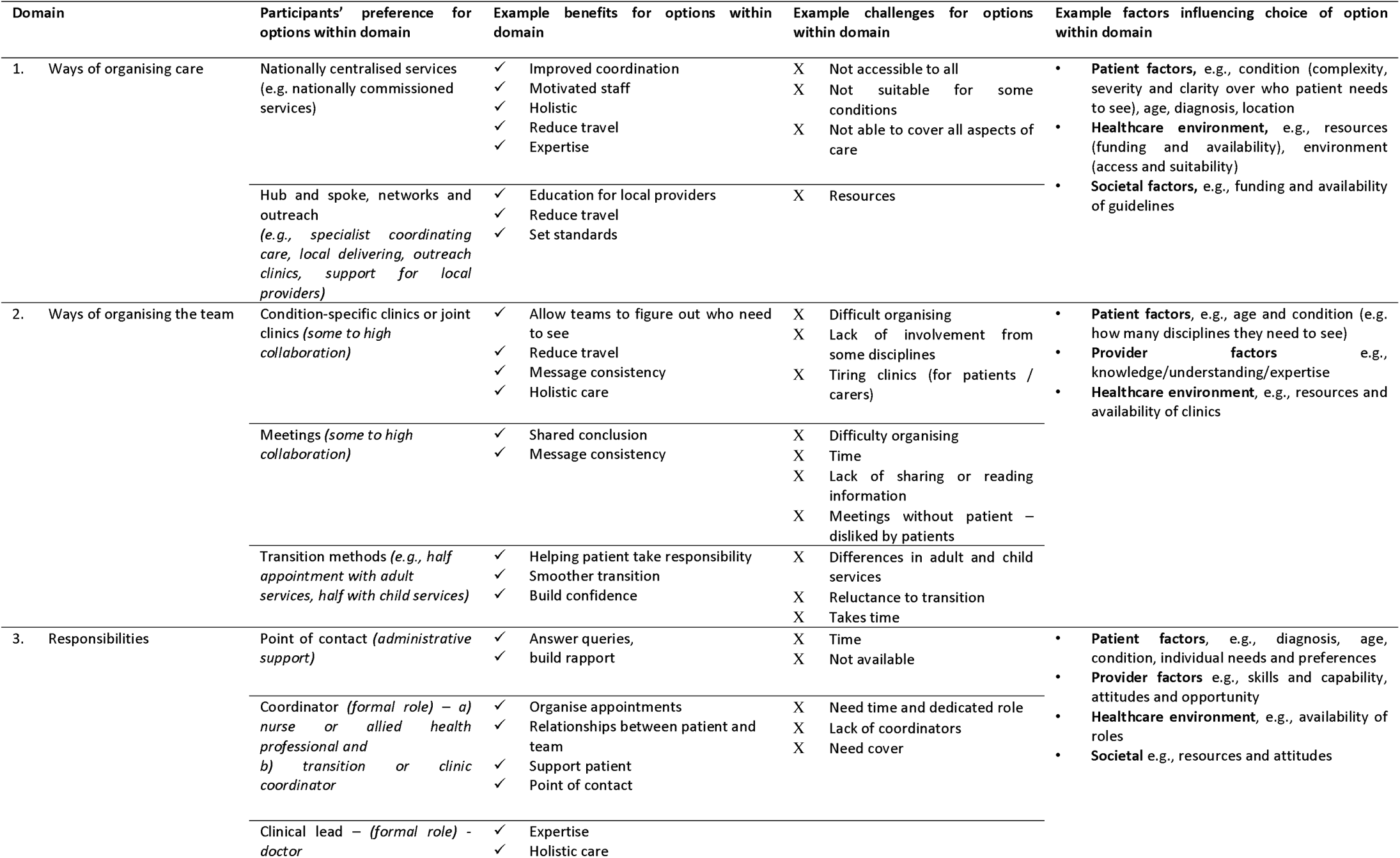
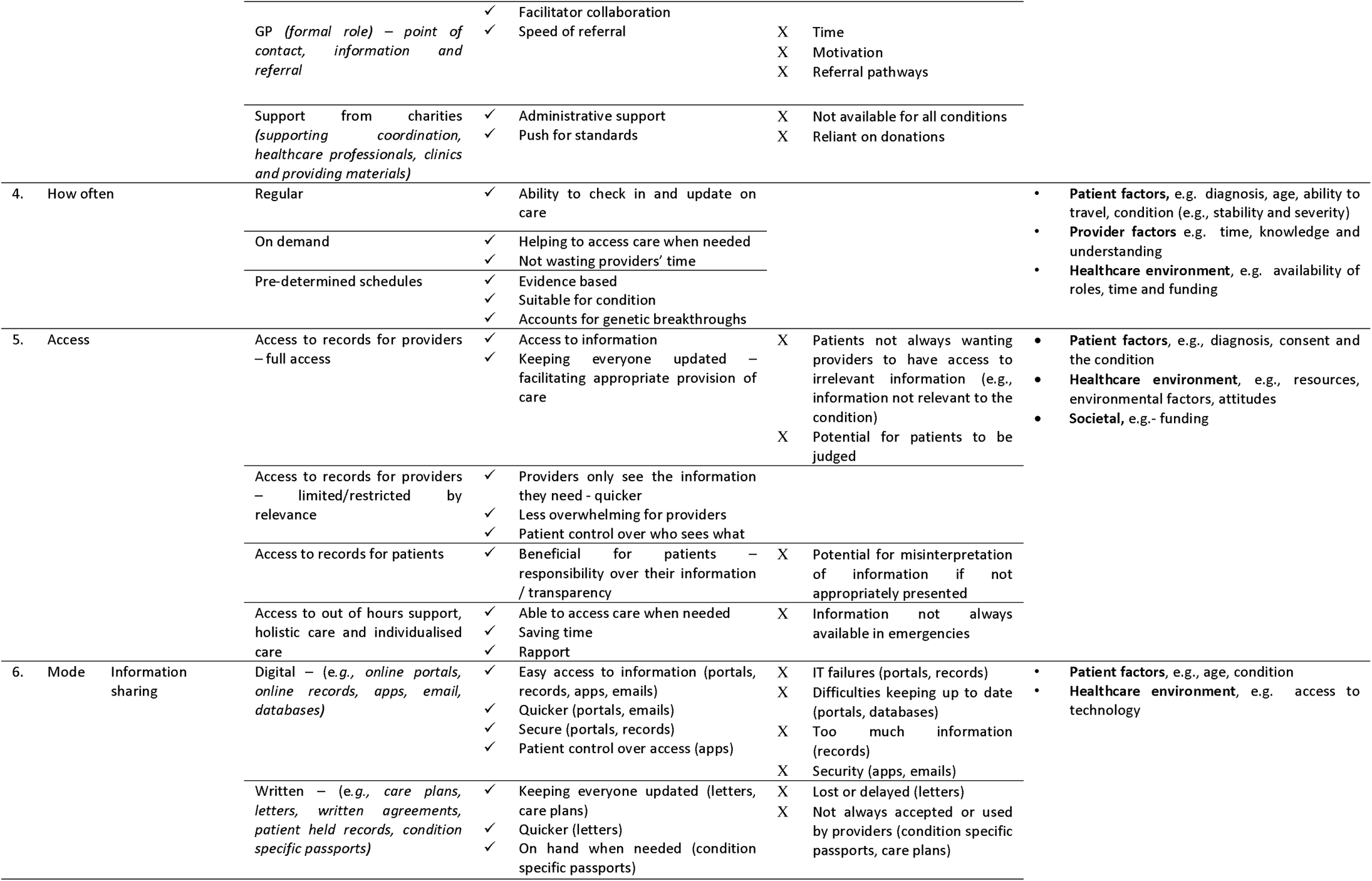
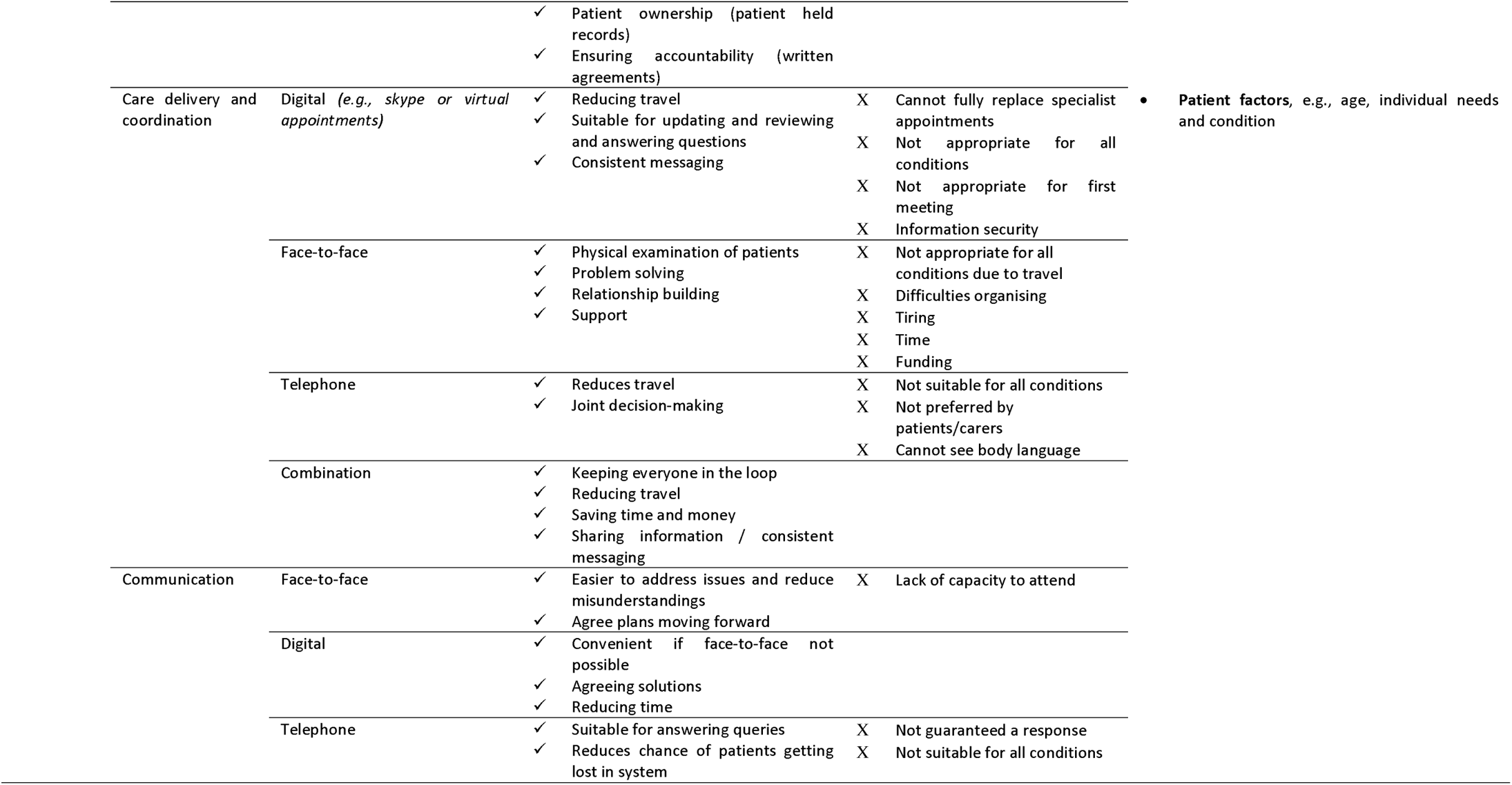
Summary of contextual findings for care coordination options, including preferences, benefits and challenges and factors influencing coordination.

#### 1. Ways of organising care

##### Which ways of organising care do stakeholders prefer?

Findings indicated that participants from all stakeholder groups may prefer nationally centralised services and hybrid models. Preferred hybrid models included specialist centres coordinating care and local services delivering care, outreach clinics and provision of support for local providers by specialist centres.

##### What are the benefits and challenges of different ways of organising care?

Nationally centralised services and hybrid models both have benefits and challenges. For example, single national centres might improve coordination and increase access to expertise. However, these services are not available for all conditions, and may not cover all aspects of care that the patient needs.

For hybrid options, these were thought to reduce travel and increase provision of education to local healthcare providers (i.e., due to specialists and local providers working together e.g. within networks or hub and spokes collaborations).

##### What factors influence the way that care is currently organised?

Many factors were perceived to influence the way care is organised, including patient factors (e.g. age, ability to travel and condition), healthcare environment factors and societal factors. Examples of healthcare factors included: availability of resources such as funding issues, availability of experts and availability of models of coordination, ease of access and suitability of the environment, and relationships between different care teams (e.g. specialist and local teams). Societal factors included funding and availability of service specifications and policies.

For example, the patient’s condition was perceived to influence how care is organised in several ways, including the nature of the condition (e.g., the complexity of the condition, whether the condition affects multiple body systems, the number of disciplines involved in a patient’s care and need for coordination across a whole spectrum of care services and not just acute medical situations). Participants also felt that specialist services (e.g. one stop shops) only work if services are able to determine exactly who a patient will need to see. Conditions that are difficult to define may not be well placed to be cared for within a specialist service. Additionally, conditions that are more stable may require less coordination (e.g., may just require a point of contact within a specialist centre).

Where the patient lives influences how care should be coordinated. Findings indicated that patients and families may fit into three groups: those who live far away from a specialist centre but can travel, those who live far away from a specialist centre but cannot travel and those who live close to a specialist centre and therefore can access it easily. Different models of care coordination may be needed for these different types of individuals/families, for example: those who live far away from the specialist centre, or are unable to travel, may require visits to specialist centres to be minimised – e.g., a greater proportion of care to be delivered locally, online or through outreach models.

#### 2. Ways of organising professionals involved in a patient’s care

##### Which ways of organising professionals do stakeholders prefer?

Findings outlined preferences for condition-specific clinics or joint clinics as opposed to individual appointments with different healthcare professionals on different days, meetings, and some transition methods to support patients (e.g. moving from child to adult services or when moving to a different location).

##### What are the benefits and challenges of different ways of organising teams?

Each of these options has benefits and challenges. For example, condition-specific clinics allow teams to figure out who patients need to see, provide access to condition-specific expertise, ensure that all those involved in a person’s care receive the same messages and may reduce travel.

However, these options are difficult to organize and multi-appointment clinics may be tiring for patients.

##### What factors influence the way that teams are organised?

Many factors were perceived to influence the type of collaboration, including patient factors (e.g., age because clinics vary for adults and children, and condition), provider factors (e.g., knowledge, understanding and whether team has multidisciplinary expertise) and healthcare environment factors (e.g., resources and availability of collaboration models such as joint clinics, MDT clinics, orientation visits, and availability of experts). For example, the nature of the condition influences collaboration as the type of clinic used depends on how multi-systemic the condition is and how many disciplines it involves. Carousel clinics or MDT clinics may only be suitable for those conditions that affect multiple body systems and MDTs may only work if there is clarity over which professionals need to be seen.

Workshop findings indicated that COVID has enabled some opportunities for collaborations between local teams and specialists (e.g. local providers dialling into multidisciplinary team meetings).

#### 3. Responsibilities

##### How would stakeholders prefer care coordination roles and responsibilities to be organised?

Findings indicated that participants from all stakeholder groups would prefer a point of contact to answer queries, a coordinator (e.g., a nurse or allied healthcare professional), a clinical lead, support from their GP and support from charities. Workshop findings highlighted the importance of charities in care coordination and also the importance of patients and carers who are often coordinating their own care.

##### What are the benefits and challenges of different ways of organising coordination roles and responsibilities?

Each of these options have benefits and challenges. For example, benefits of coordinators included helping build relationships between patients and the team and supporting patients. However, coordinators need time and a dedicated role; roles which do not consistently exist currently and require funding. People felt that clinical leads provide expertise, holistic care and facilitate collaboration between professionals. Participants felt that GPs were lacking time, sometimes motivation and clear methods to refer patients to services.

##### What factors influence who takes responsibility for coordination?

Many factors were perceived to influence who takes responsibility, including patient factors (e.g. diagnosis, age of patient, condition, and individual needs’ and preferences), provider factors (e.g. knowledge, support and education and understanding of the healthcare system, interest and motivation and time or availability of a team to work with), healthcare environment factors (e.g. resources such as availability of coordinator roles) and societal factors (e.g. availability of patient organisations, stigma and willingness to change). The patient factor that was discussed most frequently was the patient’s individual needs and preferences. For example, patient choice on who sees their records, which healthcare professionals they see, who coordinates their care and the extent to which the patient/carer are involved in coordination and meetings. Additionally, individual patient needs influence who is involved in coordination (e.g., the need for coordinated care and who is involved should be tailored and take individual family needs and ambitions into account). For example, a national care coordinator model which takes the person’s individual needs into account to determine how much contact they have with their coordinator or the level of coordination. An additional factor relating to individual needs was the patient’s ability to self-manage and coordinate their own care: some patients may be able to coordinate their own care, but others may be unable to do this and therefore need a coordinator who is more involved in their care.

Workshop findings indicated that COVID-19 may have reduced access to specialists for some participants, and limited capacity for local services and charities to support care.

#### 4. How often care appointments and coordination take place

##### Do stakeholders prefer on-demand or regular appointments?

There was less agreement between patients and carers regarding preferences for when appointments are scheduled, with some participants preferring on demand appointments for care and/or coordination, and others preferring regular appointments. However, findings from interviews, focus groups and workshops indicated that a mixture of scheduled regular appointments and on-demand appointments when needed may be preferable.

##### What are the benefits and challenges of on-demand vs regular appointments?

Some participants spoke about having preferences for on-demand appointments for care and/or coordination, as this enables them to access care when needed and not waste providers’ time. However, other participants preferred regular appointments, which would enable them to receive check-ups and update providers regularly regarding their care.

##### What factors influence whether appointments are on-demand or regular?

Many factors were perceived to influence frequency, including patient factors (e.g., diagnosis, age, ability to travel and condition – stability of the condition or the profession associated with the condition, phase, severity, individual needs, and time since treatment), provider factors (time and knowledge) and healthcare environment factors (e.g., availability of job roles, recommendations within guidelines, time within job roles and funding).

Workshop findings indicated that COVID-19 has provided some opportunities for on demand appointments (for those who have stable conditions); as long as safety nets have been put in place.

#### 5. Access to records

##### What type of access to records do stakeholders prefer?

Patients and healthcare professionals having access to records was seen as important throughout the interviews and focus groups, but there was less consensus on preferences in terms of full or filtered access. For example, for healthcare professionals, our findings show that it is important for healthcare professionals to have easy access to information and records. But the extent to which patients felt healthcare professionals should be able to access information and records varied. Some patients/carers felt that any healthcare professional should be able to access full records. Other patients/carers felt that access to records should be limited (e.g., to necessary information only).

##### What are the benefits and challenges of different types of access to records?

Perceived benefits and challenges tended to differ across patients and carers, and healthcare professionals. Some stakeholders spoke about the importance of everyone who needed access having access to records, to ensure that everyone was up to date and knew what was happening. However, some patients and carers felt that they would not want all healthcare professionals to have access to all aspects of their records (e.g., parts of their record that they considered are irrelevant) and that they would want control over who has access.

Some healthcare professionals spoke about how access to complete records can also be overwhelming and that it may be necessary to filter information by relevance.

##### What factors influence access to records?

Factors which were perceived to influence access included patient factors (e.g., diagnosis and consent), healthcare environment factors (e.g., resources, environmental factors and attitudes) and societal factors (e.g. funding).

#### 6. Modes of communication

##### Which mode of communication do stakeholders prefer?

Our participants preferred digital methods (such as online portals, records, mobile applications, emails, and databases) and written methods (such as care plans, letters, written agreements of responsibility, patient held records and condition specific passports) for information sharing. In terms of care and coordination appointments, there was less consensus (with preferences highlighted for online, face-to-face and a mixture of appointment types). For communication, participants preferred different modes depending on circumstances (e.g. telephone calls were felt to be appropriate for answering queries).

##### What are the benefits and challenges of different modes of communication?

For information sharing, digital methods were seen to provide easier and quicker access to information but were limited by IT failures and were thought to be difficult to keep up to date. Written methods were thought to keep everyone up to date and ensure accountability but may get lost or delayed.

For care and coordination appointments, each mode has benefits and challenges. Remote digital appointments may reduce travel and may be suitable for reviews and updates but cannot fully replace face-to-face appointments. Using a combination of methods was felt to keep everyone in the loop, reduce travel, save time and money, and ensure that everybody involved has the same information.

For communication, face to face methods were perceived to reduce misunderstandings and help to agree plans but were limited by availability. Remote digital methods were good for reducing time and agreeing solutions. Telephone methods were suitable for answering patient queries.

Workshop findings indicated that COVID-19 has accelerated the shift from face-to-face care to appointments involving digital or telephone methods. In some cases, COVID-19 was felt to have enabled opportunities for flexible modes that best suit the patient to be used.

##### What factors influence the mode of coordination activities?

Perceived factors influencing mode included patient factors (e.g., age, condition, and individual needs) and healthcare environment factors (e.g., access to technology).

#### Barriers and facilitators underpinning models of care coordination

Our findings also identified many barriers and facilitators underpinning these domains of care coordination. Barriers and facilitators fit within five themes (ability, attitudes, opportunity, resources and environment) (see Figure 3).

**Figure 3.**
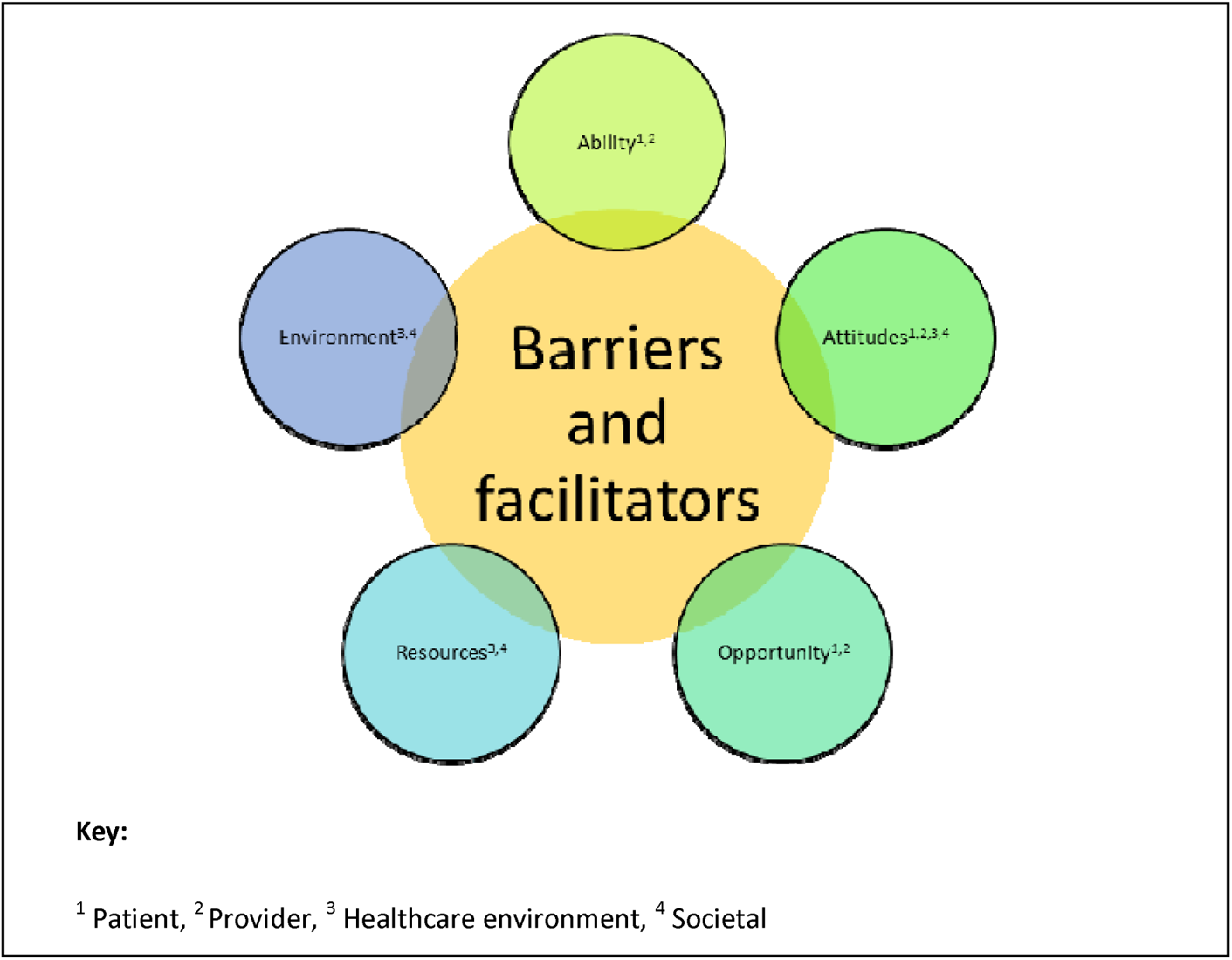
Summary of themes relating to barriers and facilitators to care coordination

Findings indicated that for patients, examples of facilitators to coordinated care were having the ability to coordinate care, self-manage their condition, having knowledge on how to coordinate care and navigate healthcare services, feeling comfortable and having a positive relationship with professionals/coordinators, and having financial ability and time to access care facilitated coordination. Alternatively, examples of barriers included: a lack of ability to self-manage and coordinate care, a lack of knowledge, anxieties and worries (e.g., not wanting to pester professionals, worries about transition and multidisciplinary clinics) and lack of finances and funding to access care.

For staff, examples of facilitators to coordinating care included: having knowledge and awareness of rare conditions, training, interest and motivation in coordinating care and taking ownership, providers’ personality and people skills, having the right mix of team members involved, having named providers and having coordinator roles. Examples of barriers included: lack of motivation and interest in coordinating care, anxieties about treating rare conditions, lack of dedicated time and other competing priorities.

In terms of the healthcare environment, examples of barriers related to resources (e.g. availability of providers, availability of technology including linked NHS IT infrastructures and funding), the environment (e.g. organisational time restraints such as ten minute appointments) and attitudes (e.g. organisational politics such as strong disciplinary boundaries and hierarchy of the NHS).

Examples of facilitators included funding and capacity, availability of facilities, cross organisational relationships and supportive organisations.

In terms of the wider society, examples of barriers included wider funding issues (e.g. care budget being split across different sectors and lack of funding for multidisciplinary work and networks), and stigma. Examples of facilitators included availability of patient groups and support from these groups.

### Aim 2. Development of hypothetical models of care coordination for rare conditions

We developed ten hypothetical models of care coordination: six for those with access to a specialist centre (models 1-6) and four for those without access to a specialist centre (models 7-10). These are summarised in Table 3 (see Appendix 6 for further details). The type of model is a function of where the patient/carer lives in relation to a specialist centre, whether the patient/carer can or wants to travel to a specialist centre, whether they have the ability (and desire) to coordinate their own care, whether they have access to a specialist centre and whether it is clear who the patient needs to see for the management of their condition. The characteristics of the models are centred around attending a specialist centre or outreach clinic, having a formalised care agreement (care plan), having a care coordinator to organise appointments (or providing a point of contact) whether there are meetings between healthcare professionals to discuss care, and the type of healthcare professional who oversees care. As noted above, the specificity of these characteristics will be determined by situation-specific factors (such as funding and staffing).

**Table 3.**
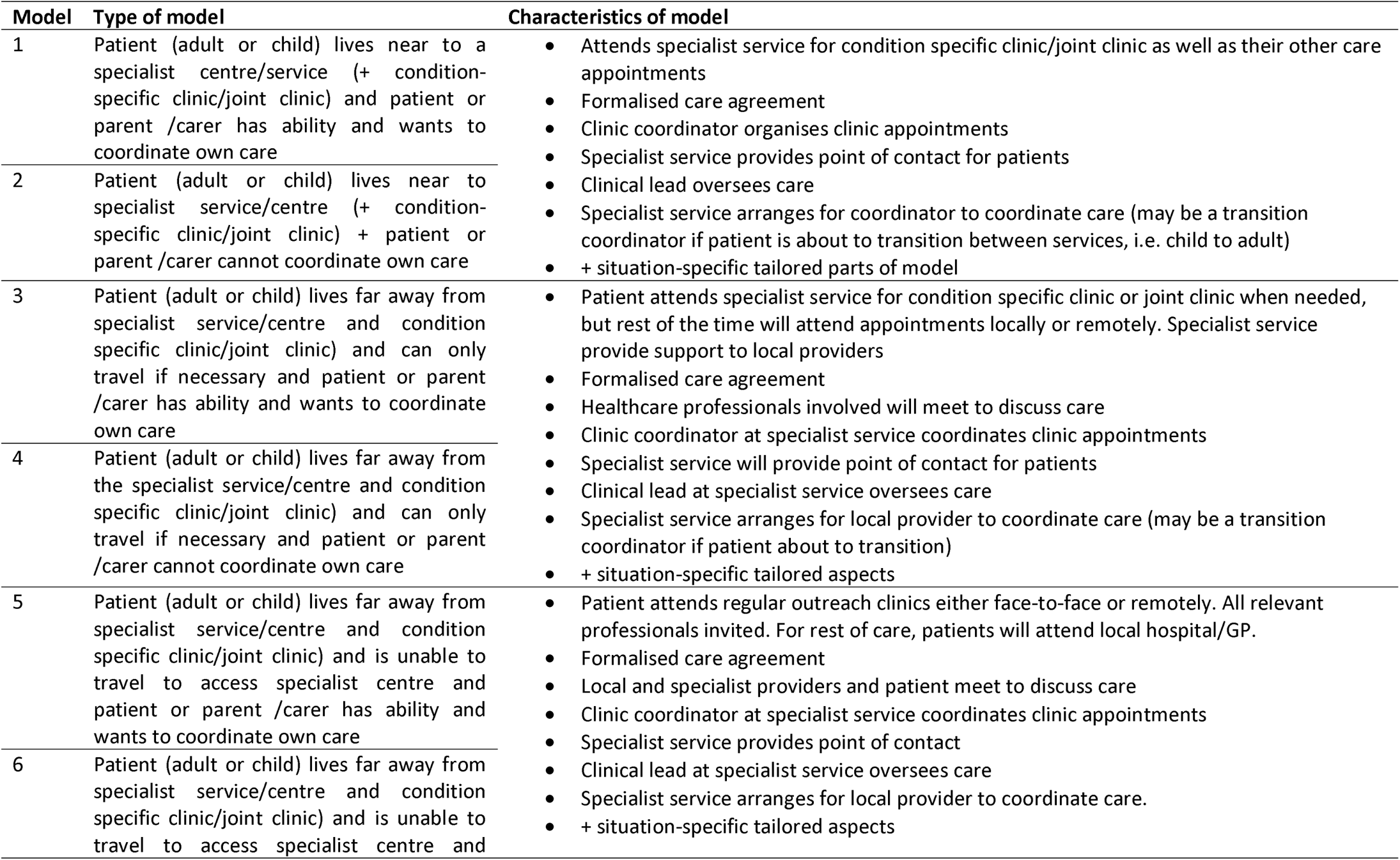
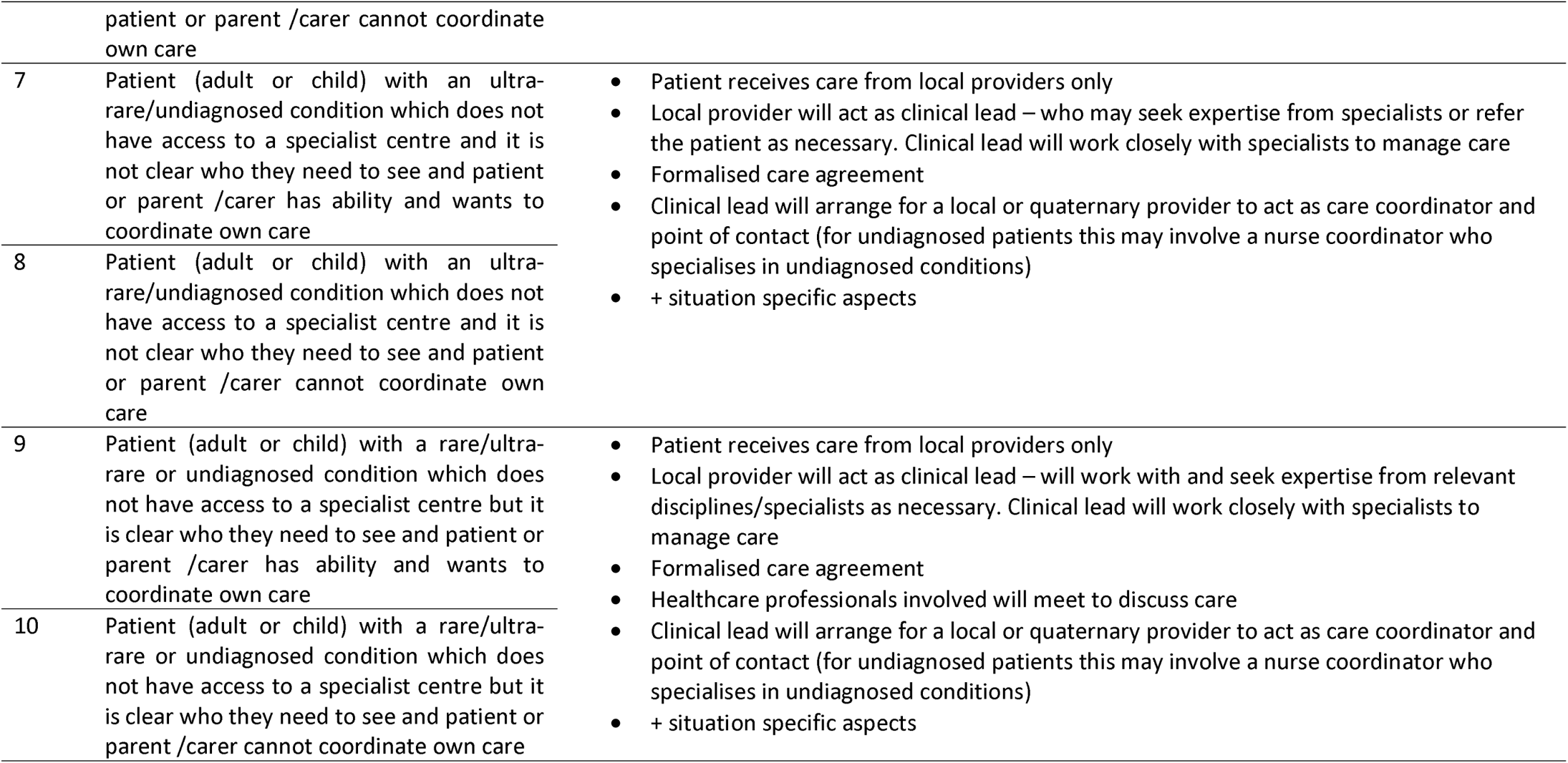
Illustrative hypothetical models of care coordination

## Discussion

### Key findings

Our findings underline that different models of care coordination for rare conditions may be appropriate in different situations. Our findings indicated that stakeholders prefer models of care that: a) are nationally centralised or a hybrid of national and local care (e.g. outreach clinics), b) involve professionals working together to provide care (e.g. in condition-specific clinics), c) include points of contact, coordinators (e.g. from nursing or allied health roles), clinical leads, GPs and charity support, d) offer regular and on demand appointments, e) provide access to records for professionals and patients, and f) use a range of digital, face-to-face and telephone modes for communication. We found a range of benefits and challenges for different types of care coordination. Our findings highlighted many factors related to the patient (e.g., condition complexity and severity, where the patient lives and whether they’re able to coordinate their own care), the healthcare professional (e.g., knowledge and time), the healthcare environment (e.g. resources) and societal factors (e.g. availability of funding) which influence the appropriateness of different care coordination options and models. We developed and refined ten illustrative models of care coordination for rare conditions, which consider different circumstances and situations, using our taxonomy [15].

### How findings relate to previous research

This research offers insight into participants’ preferences, the benefits and challenges of different models of coordination, factors influencing coordination, and barriers and facilitators to coordination in general. These findings extend previous knowledge by identifying possible situations in which different models of coordination may be appropriate. For example, previous research has highlighted that some aspects of care coordination may be necessary for rare conditions, e.g., care coordinators and specialist centres [14]. However, there has been little research on the benefits and challenges of each model for rare conditions and how they work in practice. This research extends this knowledge by outlining the factors associated with different types of coordination and using these factors to develop hypothetical models of care coordination which may be appropriate in different situations. The evidence-based process through which we have developed our models supports and extends previous research by demonstrating how we can use qualitative methods to adapt complex interventions such as care coordination to local situations, and how we can involve stakeholders in these processes [20].

Previous research has indicated that more care coordination is needed in complex situations (e.g., limited patient capacity and clinical complexity) [21]. Our findings concur with this and highlight a range of factors that need to be considered when choosing how to coordinate care, including patient, provider, environmental and societal factors. Examples of patient factors included severity and complexity of condition, where patients live, their ability to travel and ability to coordinate care. Findings therefore indicate that it is not ‘one size fits all’, and that we should develop models of care coordination that consider a range of individual, organisational and societal factors, rather than just developing different models of coordination specific to certain rare conditions. Models can then be tailored to individual situations; as with the hypothetical models proposed here. This may enable the delivery of care coordination which is equitable and family-centred, as recommended in our previous definition of care coordination [8]. Tailoring care coordination strategies to individual needs would also help to overcome some of the previously aforementioned costs associated with patients and carers coordinating their own care [7].

Our findings extend previous research by demonstrating that participants from all stakeholder groups indicated a strong preference for nationally commissioned services and hybrid models (including hub and spoke models, network models and outreach models), due to benefits which include increasing coordination, access to expertise and reducing travel. This supports previous research, which highlights the potential benefits of specialist services [22], hub and spoke models [23,24] and outreach models [25] for different health conditions. However, for rare conditions, our findings indicated that these models may not be appropriate in all situations, and in some situations patients may prefer specialist care provided locally (e.g., if they are unable to travel or do not live near to a specialist centre). Additionally, specialist services may not be appropriate for every condition. These findings highlight that different models of care coordination are needed for different types of families (i.e., those who live near to specialist centres, those who live far away but can travel, and those who live far away but cannot travel).

These findings indicate that different models of care coordination are needed to consider those who are able and want to coordinate their care and those who cannot. For example, the level/type of coordinator offered (administrative, care coordinator or clinical care coordinator) should vary depending on complexity and the patients’ ability and wish to coordinate their own care. However, findings indicate that care coordinator roles do not always exist in practice and that further resources are needed (e.g., specific roles and training pathways for coordinators). Care coordinators are not new and have previously been widely implemented for adults and children with chronic conditions and mental health conditions in other countries and in the UK [26–30]. The finding that patients/carers are sometimes unable to, or do not want to, coordinate their own care is consistent with previous research that has indicated the negative impact coordinating care can have on patients and families [7,31] and of the treatment burden more generally [1,16].

We found that each mode of communication and coordination has benefits and challenges and findings indicate that the mode of coordination should consider many factors including individual preferences and resources available. Additionally, despite the potential of remote digital methods for use in healthcare delivery [32,33], findings indicate that digital appointments must not replace face-to-face appointments completely in terms of care delivery and coordination. In person face-to-face appointments were felt to be integral, particularly at key points of the patients’ journey (e.g., initial meetings, diagnosis, potential deterioration), for certain conditions whereby face-to-face appointments are necessary, or for patients requiring more in-depth clinical care coordination due to additional difficulties. This extends previous research by highlighting the limits of remote digital methods of care delivery and coordination whilst emphasising the need to offer the option for multiple modes of delivery when coordinating care for patients with rare conditions.

### Strengths and Limitations

The findings presented in this manuscript and our resulting hypothetical models were developed from a large dataset which included participants from a wide range of roles (patients, carers, healthcare professionals, charity representatives and commissioners), who represented a wide range of rare conditions, across different locations and sectors. Therefore, whilst it is difficult to capture views from every rare condition and situation, these findings provide a clear basis for the factors that need to be considered when developing and evaluating models of care coordination.

We found that care coordination is not one size fits all and that there are many ways of coordinating care depending on individual, professional, organisational and environmental factors. Equally, we found variation in preferences for different models of care coordination and that each model had associated benefits and challenges. Therefore, we ended up developing hypothetical models instead of actual care coordination models as the findings indicated that we may not be able to fully represent all situations, domains and options of care coordination if using real life examples. However, many real-life examples of different aspects of coordination are shown in Walton et al [15].

Whilst the flow chart has facilitated the development of hypothetical models, one limitation is that it has not yet been tested or amended for use as a decision-making tool or quality improvement tool in practice. Our evidence-based process and the involvement of patients, carers, healthcare professionals, charity representatives and commissioners throughout data collection and when refining the models appears to closely align with the five phases recommended for process mapping [20]: whereby multiple perspectives from different stakeholders are sought, data are then used to develop a map, the map is validated by stakeholders and then the ideas are implemented/tested. However, we did not use the process mapping approach to develop the methods used in our study.

### Implications

Our findings indicate the need for policymakers and service commissioners to plan and develop appropriate models of care coordination which suit different needs, and which can be tailored towards different services and individuals. As we have shown in this manuscript, our taxonomy and the findings presented here can be used as a menu to help service planners think about how they develop and evaluate new models of coordination whilst considering necessary factors. For example, we present hypothetical models of care coordination that could be developed in practice and evaluated (e.g., in terms of their cost). It is hoped that the CONCORD flow chart (see Appendix 4) together with the findings can be used to facilitate decision-making processes regarding how care should be coordinated. These findings can be used by those involved in service planning, and those wanting to evaluate how care is coordinated. The findings can be used to inform which models of coordination may be suitable for use in different situations. This is particularly helpful given the complexity of care pathways and funding for rare conditions.

### Future research

Further research is needed to evaluate the implementation, effectiveness and cost-effectiveness of real-world models of coordination. To do this, further research which aims to operationalise and measure care coordination in practice is needed.

Further research would also be beneficial to test the flow chart as a decision-making tool for use by policy makers or service planners on a national and local level. If this is successful, the flow chart may have the potential to drive improvements in care coordination nationally and/or locally. For example, future research could explore the use of this tool in evaluating and informing adaptations to existing care coordination practice or informing the development and implementation of new care coordination strategies in practice. Additionally, future research could explore the use of this tool as a decision-making tool for patients and their healthcare professionals.

### Conclusions

Whilst different stakeholders have different preferences around care coordination, each type of care coordination has associated benefits and challenges. Patient/carer, provider, environmental and societal factors influence coordination. We demonstrate that it is possible to suggest hypothetical models of care coordination from the taxonomy that our findings generated. This process has highlighted that different models of care coordination may suit different circumstances, and can be used to support discussion and planning around which models may be feasible and desirable in different circumstances.

## Declarations

### Ethical approval

This study received ethical approval from the London-Surrey Borders Research Ethics Committee of the Health Research Authority (reference: 19/LO/0250).

Participants provided informed consent to participate in this study and for anonymised quotes to be published.

### Availability of data and materials

The datasets generated and/or analysed during the current study are not publicly available due to participant confidentiality but are available from the corresponding author on reasonable request.

### Author contributions

All authors have made substantial contributions to the manuscript. HW, AS, AIGR, SM, AH, LB, AGS, JK and NJF were responsible for the conception and design of the study: HW drafted the initial study protocol and all authors provided feedback. NJF, SM, AIGR and AH provided support and guidance throughout the project. LB and KLB helped with planning via the PPIAG. HW carried out interviews. HW and AS led the focus groups. HW, AIGR, SM, AH, JJ, and PLN conducted the workshops. HW, AS, and PLN organised the focus groups and workshops. HW and AS coded interview and focus group data. All authors contributed to and agreed the interpretation and analysis of the data and development of the hypothetical models. HW drafted the paper and all authors contributed to the revision of the manuscript. All authors read and approved the final manuscript.

## Acknowledgements

Thank you to the wider CONCORD team (Lyn Chitty, Emma Hudson, Sharon Parkes, Larissa Kerecuk and Christine Taylor) for providing support and feedback throughout the study. Thank you to the CONCORD Patient and Public Involvement Advisory Group for providing feedback throughout the project. Thank you to all of our participants who took part in the interviews, focus groups and workshops.

