## Appendix 1,2,3 for "Development of models of care coordination for rare conditions: A qualitative study"

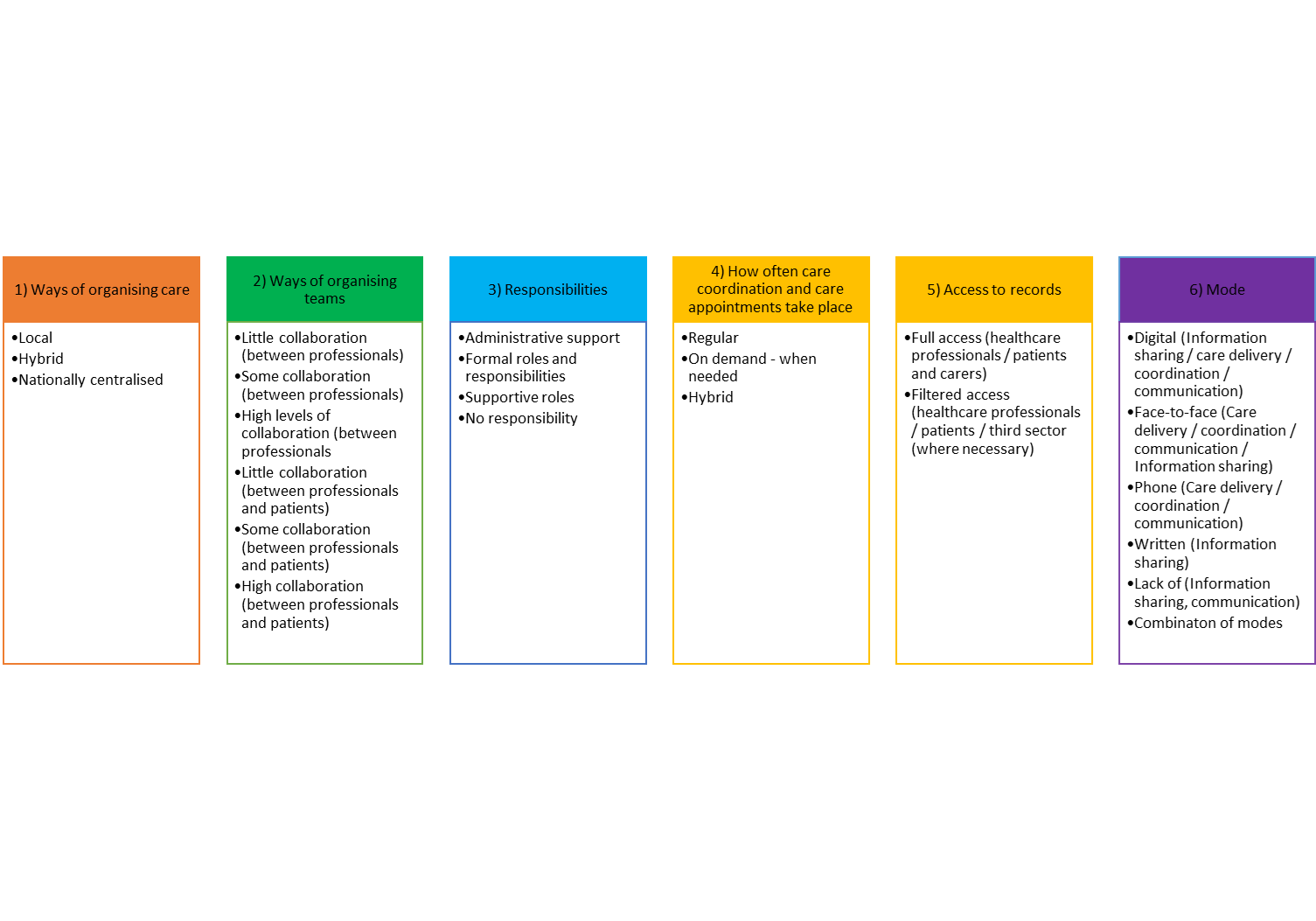
Appendix 1. A summary of the taxonomy and findings presented in [15]

**Appendix 2.** Detailed methods, (amended from [9,15; Morris et al, forthcoming – CONCORD final report])

**Design**

Our study used qualitative methods (interviews, focus groups and workshops).

Using a qualitative approach allows for a more in-depth understanding of complex phenomena (Bogardus et al, 1998; Bradley et al, 2001). Additionally, the use of qualitative methods allows for the direct involvement of those with most experience in the phenomena being studied and classified, such as patients, health care professionals and carers. This is particularly important in health care service research, in which patients, carers and health care professionals are the key stakeholders (Ferris et al, 2018). By understanding patients’, carers’ and health care professionals’ views on the organisation of care coordination for rare diseases we could improve health care services but also optimise the patient experience. It has also been proposed that qualitative studies are well suited to explore new concepts (Bradley, 2001). As coordination of care is a relatively new field, using qualitative methods will offer a rich perspective on care and stakeholders’ preferences.

This research was conducted in a two-stage process. First, interviews (n=30) and focus groups (n=4) were conducted to develop an initial taxonomy. Interviews and focus groups were felt to be appropriate methods for exploring and gathering in-depth perspectives on stakeholders experiences of coordination and the different models of care coordination that currently exist, together with preferences for potential new models of coordination. Workshops (n=2) were then conducted to refine the proposed taxonomy. Workshops were felt to be appropriate for helping to gather consensus around whether the taxonomy was appropriate and to develop recommendations to improve the taxonomy.

**Sample**

We recruited a range of patients with rare, ultra-rare or undiagnosed conditions, carers/parents, health care professionals, charity representatives and commissioners to take part in the interviews (n=30), focus groups (four groups of between six to eight participants) (Kreuger et al, 2002) and workshops (two workshops, one for patients/carers and one for professionals, of approximately 15 participants each). We originally planned for five face-to-face workshops, however we had to reduce this to two remote workshops (due to the COVID-19 pandemic). Interview and focus group participants informed the development of the taxonomy. Workshop participants informed the refinement of the taxonomy.

To take part in the study, participants needed to be aged ≥ 18 years. Children were not included due to ethical issues recruiting participants aged < 18 years. One focus group participant withdrew from the study after the focus group, thus resulting in 22 patients and carers taking part in the focus groups.

Participants were recruited using a range of methods, including email invitation, social media, via the voluntary sector and through our partnerships with four NHS sites.

As there are currently between 6,000 and 8,000 rare diseases (Rare Disease UK, 2018), it was not possible to include participants affected by every rare disease. To ensure that different models of coordinated care (including different types of care coordination and no coordination) and a wide range of experience and expertise were captured, we used purposive sampling. We sampled professionals based on their area of the UK, job role, and experience with different types of care coordination. We sampled patients and carers based on their area of the UK, condition, role, age and experience with different types of care coordination.

### Measures

To gather data to inform the development of the taxonomy, two topic guides (one for interviews and one for focus groups) were developed and used to collect data (see *Supplementary material file 2*). Questions focused on: stakeholders’ experiences of coordinated care; implications of coordinated care; preferences for key aspects of care coordination (including preferred way of coordinating care, format, access, frequency, location, information sharing and transition); benefits and challenges and factors that help and get in the way of coordination. Feedback on the topic guide was sought from the CONCORD PPIAG prior to data collection.

To gather data to refine the taxonomy, one topic guide for both workshops was developed and used to collect data (see Appendix 2). The topic guide was based around the six categories identified in the taxonomy and included prompts regarding whether the participants had feedback on the category (e.g. if we had missed anything and whether findings seemed appropriate based on participant experiences); appropriateness of options in light of the COViD-19 pandemic; and recommendations to improve the category.

### Procedure

Participants were recruited using a range of methods, including email invitation, social media, via the charity sector, and through our partnerships with four NHS sites. Potential participants were asked to contact the study researcher via email or telephone.

To ensure that a range of participants with different experiences were recruited, potential participants were asked to provide responses to eligibility questions when registering their interest. Participants were sent these eligibility questions by email. For professionals, these included: their occupation, speciality and geographical region. For patients or carers, these included whether they receive coordinated care (specialist service and who coordinates), whether they have a diagnosis, age range, ethnicity, geographical region and role. The researcher checked that participants met the eligibility criteria for the study.

Selected individuals were asked to complete consent forms (one for the researcher and one for the participant) prior to taking part in the interviews, focus groups or workshops. Participants who took part virtually or via telephone were asked to return written consent forms in advance. Participants were informed that their data would be kept confidential, fully anonymised and that they could withdraw at any time without providing a reason. Focus group participants were informed that any data collected up until the point of withdrawal would be kept due to difficulties removing individual participants from focus group data. We took steps to ensure that quotes from the participant who withdrew from the study were not included in publications. These steps included removing withdrawn quotes from the analysis spreadsheet.

To gather data to inform the development of a taxonomy, interviews with health care professionals, charity representatives and commissioners, and focus groups with patients and carers, were conducted.

One researcher conducted interviews either by telephone or face-to-face, depending on participants’ preferences. The interviews lasted approximately one hour (range: 44-74 minutes). Two researchers (HW and AS) conducted the four focus groups (one researcher facilitated, and one researcher took notes) (Kreuger, 2002). A third researcher observed one of the focus groups (EH). Two focus groups were conducted face-to-face (one in London, one in Birmingham), and two were conducted virtually using Skype for Business. Focus groups were up to three hours in length (including a break) (range: 149-154 minutes). Interviews and focus groups were digitally recorded using an encrypted dictaphone (with consent from participants) and professionally transcribed. Transcripts were checked for accuracy and fully anonymised (including names and places). Data were stored in the UCL Data Safe Haven (a secure electronic environment, certified to ISO27001 information security standard and conforms to the NHS Information Governance Toolkit) and coded using NVivo 12.

To refine the taxonomy, workshops were conducted virtually. Workshop participants were sent a brief 15-minute video prior to the workshop which outlined the findings of the taxonomy (including each domain and the options within each domain). The presentation also covered qualifier findings including preferences, barriers/facilitators, factors influencing coordination and benefits/challenges of different options. During the workshops, participants were given an introduction to the workshop before being split into three breakout groups. Each breakout group had one facilitator (HW, EH, AIGR) and one note taker (JJ, SM, AH). During the breakout groups, facilitators encouraged the small groups to discuss each of the six domains in the taxonomy. Facilitators prompted about whether participants had feedback on the category (i.e. whether anything had been missed, whether findings seemed appropriate based on participant experiences), appropriateness of options in light of the pandemic and recommendations to improve the category. After the breakout groups, participants reconvened in the main group and each group provided feedback on their discussions. Workshops were recorded using an encrypted dictaphone. Notes were checked for thoroughness, and summarised prior to being sent to a graphic facilitator (New Possibilities, Birmingham, UK) to create a graphical representation of the findings.

**Appendix 3.** Topic guides for interviews, focus groups and workshops

**Interviews**

| **Interview questions** | **Prompts** |
| --- | --- |
| 1. Please tell me a bit about yourself. | - Job role - Experience of rare or ultra-rare diseases - Experience providing or being involved in the provision of coordinated care |
| 2. What types of coordinated care are you aware of? | - Models of coordination - Coordination addressing different types of transitions - How is this model similar to other models of coordinated care? - How is this model different from other models of coordinated care? |
| 3. What are the implications of coordinated care/lack of coordinated care? | - Positive - Negative - Please could you give an example of where things went well or didn’t go well? |
| 4. What type of coordinated care would you like to be delivered? | - What would this involve? - How would this be done? - What would this look like if it were successfully used in practice? - Please could you give an example of where this type of coordination went well or didn’t go well? - Please can you tell me a bit about why? |
| 5. What would be your preferred way for patients and family members to access coordinated care? | - What would this involve? - How would this be done? - What would this look like if it were successfully used in practice? - Please could you give an example of where this went well or didn’t go well? - Please can you tell me a bit about why? |
| 6. What would be your preferred format of coordinated care? | - What would this involve? - How would this be done? - What would this look like if it were successfully used in practice? - Please could you give an example of where this went well or didn’t go well? - Please can you tell me a bit about why? |
| 7. What would be your preference on how often patients receive coordinated care? | - What would this involve? - How would this be done? - Would this differ depending on the type of coordinated care provided? - What would this look like if it were successfully used in practice? - Please could you give an example of where this went well or didn’t go well? - Please can you tell me a bit about why? |
| 8. What would be your preferences on where care coordination is provided? (where appropriate) | - What would this involve? - How would this be done? - What would this look like if it were successfully used in practice? - Please could you give an example of where this went well or didn’t go well? - Please can you tell me a bit about why? |
| 9. What would be your preferences on how information would be shared between healthcare providers, patients and carers and local services? (where appropriate) | - What would this involve? - How would this be done? - What would this look like if it were successfully used in practice? - Please could you give an example of where this went well or didn’t go well? - Please can you tell me a bit about why? |
| 10. What would your preferred method of transition (movement) between services (e.g. child to adult) be for patients? | - What would this involve? - How would this be done? - What would this look like if it were successfully delivered in practice? - Please could you give an example of where this went well or didn’t go well? - Please can you tell me a bit about why? |
| 11. How do you think that patients and carers would like their care to be coordinated? | - Please can you tell me a bit about why? |
| 12. What things need to be taken into account when deciding how best to coordinate care? | - E.g. different conditions / different age ranges - Please can you tell me a bit about why? |
| 13. What factors help to provide coordinated care? | - Locally  - Nationally  - How did/might they help?  - Have you been involved in initiatives to improve coordination previously? If so, what changes helped to improve care coordination? |
| 14. What factors get in the way of providing coordinated care? | - Locally  -Nationally  - How did/might they get in the way?  - How do you overcome these problems? |
| 15. Is there anything else that you would like to say about what we have talked about? | - Is there anything else that you would like to mention? - Are there any important issues that have not been raised? |

**Focus group**

| **Structure** | **Prompts (if needed)** |
| --- | --- |
| 16. Let’s begin. Let’s find out some more about each other by going around the table. Please tell us your name, whether you are a patient or parent/carer and where you are from |  |
| 17. Please tell us about your experiences of coordinated care (approx. 2 mins each) | - E.g. fully coordinated care, some coordinated care, no coordinated care   *<After each person>*   - Which aspects of your care that were coordinated worked well? - Which aspects of your care could be coordinated better? |
| 18. What are the implications of having/not having coordinated care? | - Positive - Negative - Please could you give an example of where things went well or didn’t go well? - E.g. number of clinics people attend/how far they have to travel - E.g. psychological, clinical, medical, social, financial implications |
| 19. **Thinking about the different types of coordinated care, please identify your preferred choices for the following aspects of coordinated care:**     - How would you like your care to be coordinated?     - Which aspects of care would care coordination matter most to you?     - Which aspects of care would care coordination not matter to you?     - How would you like to access coordinated care     - How would you like to communicate with other people involved in care coordination?     - How often would you want to receive coordinated care?     - Where would you like care to be coordinated?     - How would you like information to be shared between healthcare providers, patients and carers, and local services?     - What is your preferred method of transition (or movement) across services? | *<Go through each of the questions one by one and prompt the following questions>:*   - Please can you tell me a bit about why this is your preference? - What are other people’s views on this? - How could this be done? - What would this look like if it were successfully used in practice? - Would your preferences change over time? Why? |
| 20. What factors affect your access to coordinated care? | - Locally - Nationally   - How did they help?  - How did they get in the way? |
| 7. What choice do you have in terms of the care coordination that you receive? | - How do you find this? - What choices would you like to make in relation to care coordination? - What could be improved? |
| 8. Is there anything else that you would like to say about what we have talked about? | - Is there anything else that you would like to mention? - Are there any important issues that have not been raised? |

**Workshops**

| Time | Tasks/sessions |
| --- | --- |
| 10 minutes | Introduction to workshop and ground rules & brief intro to participants & brief recap of video/introduce task |
| 40 minutes | Group discussion on taxonomy (domains and characteristics) – go through each domain answering the following questions:   - What’s good about this domain and the characteristics within it? (10 mins) - What needs improving? (10 mins) - Appropriateness of characteristics within this domain in relation to use during current COVID situation? (10 mins) - Recommendations to improve domain/characteristics? (10 mins)   If time left – could also ask similar questions about the models |
| 10 minutes | Development of recommendations to improve taxonomy and models (summary from discussion and any other thoughts?) |
| 5 minutes | Introduce optional activity for after workshop (if they would like to they can provide feedback on models using the following questions:   - What’s good about the model? - What needs improving? - Appropriateness of model in relation to use during current COVID situation? - Recommendations to improve model?) |
| 5 minutes | Questions and summary/debrief |
