## Appendix 4 for "Development of models of care coordination for rare conditions: A qualitative study"

### CONCORD Flow Chart demonstrating how taxonomy options can be used to develop models of care coordination

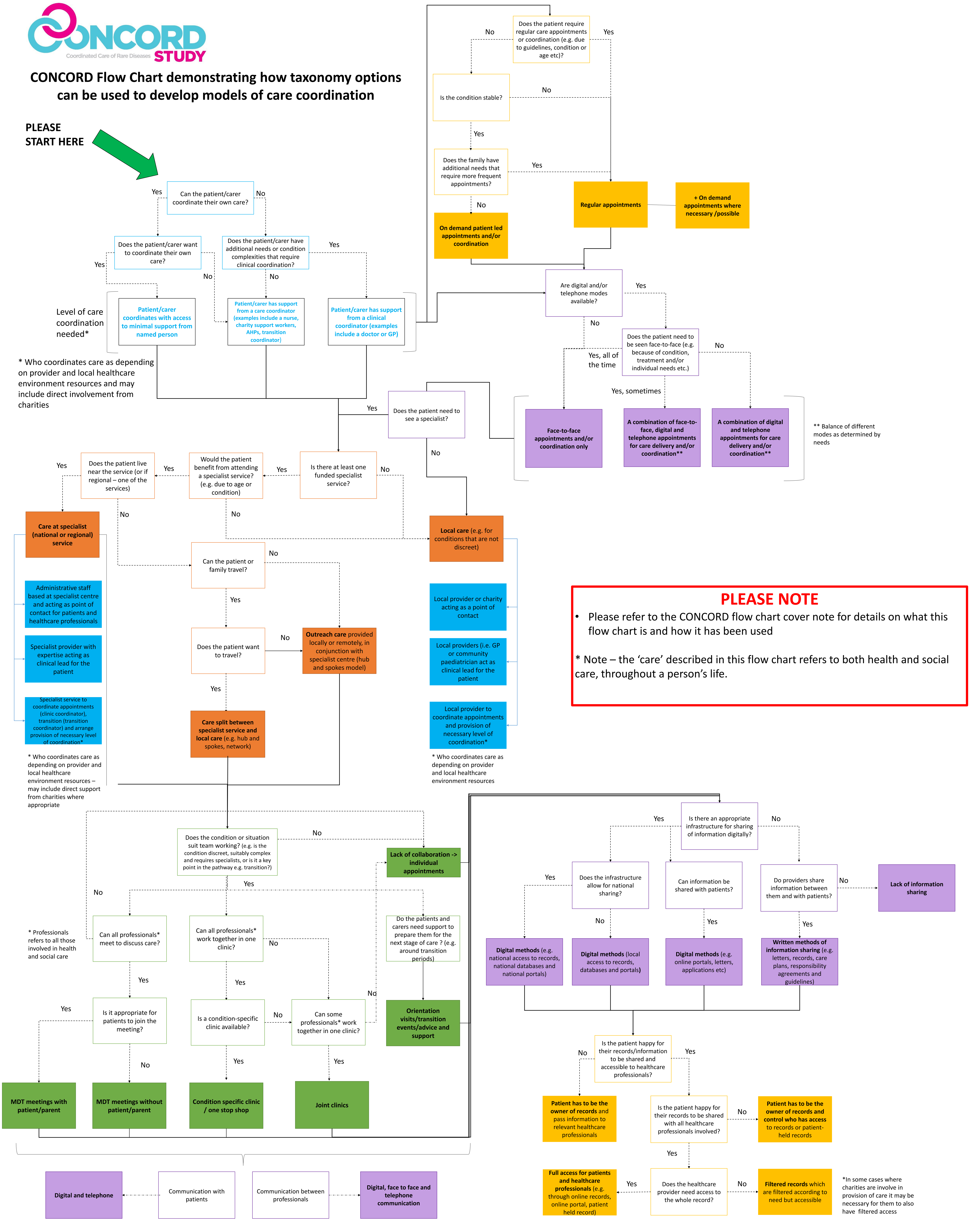


#### PLEASE NOTE

- Please refer to the CONCORD flow chart cover note for details on what this flow chart is and how it has been used

* Note – the 'care' described in this flow chart refers to both health and social care, throughout a person's life.
