## Appendix 5 for "Development of models of care coordination for rare conditions: A qualitative study"

**Cover note for CONCORD flow chart demonstrating how taxonomy options can be used to develop models of care coordination**

### How has this flow chart been developed?

This flow chart has been developed from the qualitative CONCORD study which aimed to develop a taxonomy of different ways of coordinating care for rare conditions.

These findings are based on 30 interviews with healthcare providers, commissioners and charity representatives, and four focus groups with patients and carers with experience of rare, ultra-rare or undiagnosed conditions. The decisions within this flow chart are based on findings of different ways of coordinating care and  stakeholder's preferences, benefits/challenges of different models, factors influencing coordination and barriers/facilitators (as presented in the CONCORD workshops).

**What is the CONCORD flow chart?**

- The CONCORD flow chart is a visual representation of the findings from the CONCORD taxonomy study, and has been used by the CONCORD researchers to visualise how different ways of coordinating care may be used in certain situations. The flow chart could also potentially be used by commissioners/clinicians when thinking about how to coordinate care for people living with rare conditions.

**How has the CONCORD flow chart been used so far?**

- This flow chart has been used to develop potential hypothetical models of care coordination which take into account different situations. (Please see handout: 'Hypothetical models of care coordination_06012021').
- Whilst this flow chart has been used to develop hypothetical models, it has not yet been tested as such.

**How to use the CONCORD flow chart**

- There is flexibility needed in interpreting the CONCORD flow chart.
- When using the flow chart, it may be helpful to start in the top left hand corner, with the question 'Can the patient/carer coordinate their own care?'
- Within each decision box (the boxes that are fully shaded), there are multiple options that may be suitable (e.g. the type of technology used, type of mode, or who coordinates care). Additionally, in some cases 'sometimes' may be appropriate, or individuals' needs and situations may change.
- This flow chart is therefore a guide and may not account for all possible options. The flow chart may instead support discussion and thinking around which models may suit different situations.
- Further work would be necessary to ensure that this would be fit for purpose if it were to be used as a decisional tool.

**Where to start?**

- The different colour boxes represent the six domains of the CONCORD taxonomy
  - Orange = ways of organising care
  - Green = ways of organising teams
  - Blue = responsibilities
  - Yellow = access to records and frequency of appointments
  - Purple = mode
- The domains can be thought about it any order. However, as all six domains have been found to be important in coordinating care, it is important that each domain is considered when thinking about your options.
