## Appendix 6 for "Development of models of care coordination for rare conditions: A qualitative study"

06/01/2021

### Hypothetical models of care coordination

In this document, we have outlined some potential hypothetical models of care coordination.

These hypothetical models could be used by researchers to evaluate how much different models could cost, or the effectiveness of different models. These hypothetical models could also potentially be used by commissioners/clinicians when thinking about how to coordinate care for people living with rare conditions.

**How have these hypothetical models been developed?**

These hypothetical models have been developed from the qualitative CONCORD study which aimed to develop a taxonomy of different ways of coordinating care for rare conditions.

These findings are based on 30 interviews with healthcare providers, commissioners and charity representatives, and four focus groups with patients and carers with experience of rare, ultra-rare or undiagnosed conditions. These models are based on findings of different ways of coordinating care and stakeholder's preferences, benefits/challenges of different models, factors influencing coordination and barriers/facilitators (as presented in the CONCORD workshops). We presented these models to workshop participants and have made amendments to the models to take feedback into account.

Please see the following pages for examples for each of the hypothetical models.

Within these hypothetical models, we have tried to consider different scenarios into account:

- Where the patient and carer lives in relation to a specialist centre
- Whether the patient and carer can or wants to travel to a specialist centre
- Whether the patient and carer has the ability (and wants) to coordinate their own care
- Whether the patient and carer has access to a specialist centre
- Whether it is clear who the patient/carer needs to see

There are ten proposed models in total:

#### Models for conditions that have access to specialist centres/services

- Patient who lives nearby to specialist centre (only relevant for those with rare/ultra-rare conditions that have specialist centres)
    - Patient/carer has ability to coordinate own care (Model 1)
    - Patient/carer cannot coordinate own care (Model 2)
- Patient who lives far away from specialist centre but can travel (only relevant for those with rare/ultra-rare conditions that have specialist centres
    - Patient/carer has ability to coordinate own care (Model 3)
    - Patient/carer cannot coordinate own care (Model 4)
- Patient who lives far away from specialist centre but cannot travel
    - Patient/carer has ability to coordinate own care (Model 5)

  - Patient/carer cannot coordinate own care (Model 6)

#### Models for conditions that do not have access to specialist centres/services

- Patient who has an ultra-rare or undiagnosed condition which does not have a specialist centre and it is not clear who they need to see
  - Patient/carer has ability to coordinate own care (Model 7)
  - Patient/carer cannot coordinate own care (Model 8)
- Patient who has a rare, ultra-rare or undiagnosed condition which does not have a specialist centre but it is clear who they need to see
  - Patient/carer has ability to coordinate own care (Model 9)
  - Patient/carer cannot coordinate own care (Model 10)

##### Additional situation-specific decisions to be considered within each model

Within all of these models, there are situation-specific, tailored decisions to be made between the provider and patient around:

- **Level of coordinator support** (depending on condition complexity and ability to coordinate own care) e.g. minimal/moderate/high
- **Who the coordinator and clinical lead is (**who coordinator is depends on resources)
- **Who is involved in MDT meetings (**including patient and carer involvement)
- **The extent to which face-to-face/digital/phone modes are used for information sharing, communication, care delivery and coordination** (e.g. depends on how often patient's need to be seen face-to-face and technology available)
- **The extent to which information is shared** (depending on IT infrastructure) (digital, written or verbal)
- **The extent to which providers have access to records** (full/restricted/none)
- **How often care coordination and care appointments are needed** (e.g. on demand or regular)
- **Transition needs** – for those transitioning from child to adult, or from one location to another (e.g. transition coordinator, visits to the adult clinic and joint clinic)

**Key principles to base decisions on**

These decisions should be based on things like availability of resources, the environment, and patient factors and provider factors. Some examples are given in the models below but these are only examples and should be considered based on the individual context and situation.

### Models of care coordination for conditions that have access to specialist centres

**Model 1. Patient (adult or child) lives near to specialist service/centre (+condition-specific clinic/joint clinic) + patient/carer has ability and wants to coordinate own care**

- The patient will attend a specialist service (as required) for a condition specific clinic (where they will see multiple healthcare professionals in one day) or joint clinic (e.g. couple of professionals in one day or between their paediatric and adult providers to support transition), and the rest of their care appointments.
- A formalised care agreement outlining responsibilities, care and team working should be developed (E.g. contracts and access to records).
- The clinic coordinator based at the specialist service coordinates the clinic appointments.
- The specialist service will provide a point of contact for patients (e.g. administrator or nurse email system/phone system) (including out of hours contact if needed)
- A clinical lead based at the specialist service who has expertise in the condition oversees care.
- The specialist service arranges for a coordinator (E.g. community matron or nurse/allied healthcare professional based at specialist hospital based on resources) to act as the care coordinator for the patient. If the patient is about to transition from child to adult care, then a transition coordinator may be suitable for this role

**Examples of situation-specific, tailored parts of model**

*(depending on patient/provider/environmental context)*

- Patient mostly coordinates care themselves but has access to a minimal level of support from care coordinator (e.g. digital coordination appointments with a nurse if needed)
- The patient requires ongoing care appointments, but the patient & healthcare professionals together decide that they need care coordinator appointments less regularly (e.g. every 12 months) just to check in
- The hospital's infrastructure will be used to share electronic records internally. Letters will be sent to local providers and the patients to keep them updated. All healthcare providers involved in a patient's care will have restricted access to the records (seeing the information that is relevant to them).
- Mode of appointments, communication and coordination (digital, remote, phone to phone), may vary depending on factors such as situation, resources and patient preferences
- Patients may be put in contact with patient support groups where applicable

**Model 2. Patient (adult or child) lives near to specialist service/centre (+condition-specific clinic/joint clinic) + patient/carer cannot coordinate own care**

- The patient will attend a specialist service (as required) for a condition specific clinic (where they will see multiple healthcare professionals in one day) or joint clinics (e.g. couple of professionals in one day or between their paediatric and adult providers to support transition), and the rest of their care appointments.
- A formalised care agreement outlining responsibilities, care and team working should be developed (E.g. contracts and access to records).
- The clinic coordinator based at the specialist service coordinates the clinic appointments.
- The specialist service will provide a point of contact for patients (e.g. administrator or nurse email system/phone system) (including out of hours contact if needed)
- A clinical lead based at the specialist service who has expertise in the condition oversees care.
- The specialist service arranges for a coordinator (E.g. community matron or nurse/allied health care professional based at specialist hospital based on resources) to act as the care coordinator for the patient. If the patient is about to transition from child to adult care, then a transition coordinator may be suitable for this role

**Examples of situation-specific, tailored parts of model**

*(depending on patient/provider/environmental context)*

- Patient requires a high level of support from care coordinator to coordinate treatments and care (e.g. face-to-face appointments with a doctor)
- The patient requires ongoing care appointments, and ongoing coordination appointments, due to their complex care needs
- The hospital's infrastructure will be used to share electronic records internally. Letters will be sent to local providers and the patients to keep them updated. All healthcare providers involved in a patient's care will have full access to the records (seeing the information that is relevant to them).
- Mode of appointments, communication and coordination (digital, remote, phone to phone), may vary depending on factors such as situation, resources and patient preferences
- Patients may be put in contact with patient support groups where applicable

**Model 3. Patient (adult or child) lives far away from specialist service/centre (& condition-specific clinic/joint clinic) and can only travel if necessary + patient/carer has ability and wants to coordinate own care**

- The patient will attend a specialist service for a condition specific clinic (where they will see multiple healthcare professionals in one day) or joint clinic (couple of professionals in one day or between their paediatric and adult providers to support transition) when needed, but the rest of the time will either attend appointments via remote virtual appointment with the specialist service or via a face-to-face local appointment with local healthcare professionals to minimise travelling. The specialist service will provide education and support to local providers as part of a hub and spokes network.
- A formalised care agreement outlining responsibilities, care and team working should be developed (E.g. contracts and access to records).
- The healthcare professionals involved in the patient's care will meet to discuss their care (specialist providers may meet to face to face, with the local providers and coordinator dialling in).
- The clinic coordinator based at the specialist service coordinates the clinic appointments.
- The specialist service will provide a point of contact for patients (e.g. administrator or nurse email system/phone system) (including out of hours contact if needed)
- A clinical lead based at the specialist service who has expertise in the condition oversees care.
- The specialist service arranges for a local provider (E.g. community matron or nurse at local hospital based on resources) to act as the care coordinator for the patient. If the patient is about to transition from child to adult care, then a transition coordinator may be suitable for this role.

**Examples of situation-specific, tailored parts of model**

*(depending on patient/provider/environmental context)*

- Patient mostly coordinates care themselves but has access to a minimal level of support from care coordinator (e.g. digital coordination appointments with a nurse if needed)
- The patient requires on demand care appointments and coordination appointments
- The patient would only like certain healthcare professionals to have access to their records and would like to control access. As it is not yet possible to do this with electronic records, the patient will have to hold their own patient held records and give access to providers as and when needed.
- Mode of appointments, communication and coordination (digital, remote, phone to phone), may vary depending on factors such as situation, resources and patient preferences
- Patients may be put in contact with patient support groups where applicable

**Model 4. Patient (adult or child) lives far away from the specialist service/centre (& condition-specific clinic/joint clinic) and can only travel if necessary + patient/carer cannot coordinate own care**

- The patient will attend a specialist service for a condition specific clinic (where they will see multiple healthcare professionals in one day) or joint clinic (couple of professionals in one day or between their paediatric and adult providers to support transition) when needed, but the rest of the time will either attend appointments via remote virtual appointment with the specialist service or via a face-to-face local appointment with local healthcare professionals to minimise travelling. The specialist service will provide education and support to local providers as part of a hub and spokes network.
- A formalised care agreement outlining responsibilities, care and team working should be developed (E.g. contracts and access to records).
- The healthcare professionals involved in the patient's care will meet to discuss their care (specialist providers may meet to face to face, with the local providers and coordinator dialling in).
- The clinic coordinator based at the specialist service coordinates the clinic appointments.
- The specialist service will provide a point of contact for patients (e.g. administrator or nurse email system/phone system) (including out of hours contact if needed)
- A clinical lead based at the specialist service who has expertise in the condition oversees care.
- The specialist service arranges for a local provider (E.g. community matron or nurse at local hospital based on resources) to act as the care coordinator for the patient. If the patient is about to transition from child to adult care, then a transition coordinator may be suitable for this role.

**Examples of situation-specific, tailored parts of model**
*(depending on patient/provider/environmental context)*

- Patient requires a moderate level of support from care coordinator (e.g. face-to-face appointments with a nurse).
- The patient requires ongoing care appointments, but the patient & healthcare professionals together decide that they need care coordinator appointments less regularly (every 6 months).
- The hospital's infrastructure will be used to share electronic records internally. Letters will be sent to local providers and the patients to keep them updated. All healthcare providers involved in a patients care will have restricted access to the records (seeing the information that is relevant to them).
- Mode of appointments, communication and coordination (digital, remote, phone to phone), may vary depending on factors such as situation, resources and patient preferences.
- Patients may be put in contact with patient support groups where applicable.

**Model 5. Patient (adult or child) lives far away from the specialist service/centre (& condition-specific clinic/joint clinic) and is unable to travel to access specialist centre + patient/carer has the ability and wants to coordinate care**

- The patient will attend regular outreach clinics either face-to-face (provided by the specialist centre in the patient's local area) or remotely (e.g. virtually attending from GP appointment). All professionals that would be included in a multidisciplinary team are invited (some may be unable to attend). For example, a multidisciplinary outreach clinic may be delivered in the patient's local area, by the doctor with expertise from the specialist service together with a nurse and one allied healthcare professional from the specialist centre, and the patient's local GP and coordinator. (Note. How many professionals need to be involved in the outreach clinic depends, therefore, in some cases the outreach clinic may be delivered by one healthcare professional). For the rest of their care, the patient's will attend their local hospital or GP.
- A formalised care agreement outlining responsibilities, care and team working should be developed (E.g. contracts and access to records).
- The local and specialist providers together with the patient will meet to discuss their care (as part of the outreach clinic).
- The clinic coordinator based at the specialist service coordinates the clinic appointments.
- The specialist service will provide a point of contact for patients (e.g. administrator or nurse email system/phone system) (including out of hours contacts if needed)
- A clinical lead based at the specialist service who has expertise in the condition oversees care.
- The specialist service arranges for a local provider (E.g. community matron or nurse at local hospital based on resources) to act as the care coordinator for the patient.

**Examples of situation-specific, tailored parts of model**

*(depending on patient/provider/environmental context)*

- Patient mostly coordinates care themselves but has access to a minimal level of support from care coordinator (e.g. digital coordination appointments with a nurse if needed)
- The patient requires regular care appointments and on demand coordination appointments.
- Remote MDT clinic suitable for patient and MDT.
- The patient would only like certain healthcare professionals to have access to their records and would like to control access. As it is not yet possible to do this with electronic records, the patient will have to hold their own patient held records and give access to providers as and when needed.
- Mode of appointments, communication and coordination (digital, remote, phone to phone), may vary depending on factors such as situation, resources and patient preferences.
- Patients may be put in contact with patient support groups where applicable.

**Model 6. Patient (adult or child) lives far away from the specialist service/centre (& condition-specific clinic/joint clinic) and is unable to travel to access specialist centre + patient/carer cannot coordinate own care**

- The patient will attend regular outreach clinics either face-to-face (provided by the specialist centre in the patient's local area) or remotely (e.g. virtually attending from GP appointment). All professionals that would be included in a multidisciplinary team are invited (some may be unable to attend). For example, a multidisciplinary outreach clinic may be delivered in the patient's local area, by the doctor with expertise from the specialist service together with a nurse and one allied healthcare professional from the specialist centre, and the patient's local GP and coordinator. (Note. how many professionals need to be involved in the outreach clinic depends, therefore, in some cases the outreach clinic may be delivered by one healthcare professional). For the rest of their care, the patient's will attend their local hospital or GP.
- A formalised care agreement outlining responsibilities, care and team working should be developed (E.g. contracts and access to records).
- The healthcare professionals involved in the patient's care will meet to discuss their care (as part of the outreach clinic) and provide support to the local providers.
- The clinic coordinator based at the specialist service coordinates the clinic appointments.
- The specialist service will provide a point of contact for patients (e.g. administrator or nurse email system/phone system) (including out of hours contact if needed).
- A clinical lead based at the specialist service who has expertise in the condition oversees care.
- The specialist service arranges for a local provider (E.g. community matron or nurse at local hospital based on resources) to act as the care coordinator for the patient.

**Examples of situation-specific, tailored parts of model**

*(depending on patient/provider/environmental context*

- Patient requires a moderate level of support from care coordinator (e.g. face-to-face appointments with a nurse).
- The patient requires regular care appointments, and regular coordination appointments.
- Face-to-face MDT clinic needed in local area for patient and MDT.
- The specialist service and GP have full access to the patient's online records. Letters are sent to local healthcare providers and to the patient to update them.
- Mode of appointments, communication and coordination (digital, remote, phone to phone), may vary depending on factors such as situation, resources and patient preferences.
- Patients may be put in contact with patient support groups where applicable.

### Models of care coordination for conditions that do not have access to specialist centres

**Model 7. Patient (adult or child) with an ultra-rare/undiagnosed condition which does not have access to a specialist centre and it is not clear who they need to see + patient/carer has ability and wants to coordinate own care**

- The patient will receive care from their local providers only, as there is no specialist service for them to attend.
- A local provider (GP for adults, community paediatrician for children) will act as clinical lead and will oversee the patient's care. The clinical lead may seek expertise from specialists or refer the patient to quaternary hospitals e.g. geneticists etc. The clinical lead will work closely with these specialists/quaternary services to manage care (e.g. some options of this include GPs in clinical genetics depts. Or an expansion of the role of geneticists).
- A formalised care agreement outlining responsibilities, care and team working should be developed (E.g. contracts and access to records).
- The clinical lead will arrange for a local or quaternary provider (E.g. a community matron or nurse at local hospital based on resources) to act as the care coordinator and point of contact for the patient. For undiagnosed patients, this may involve a nurse coordinator who specialises in undiagnosed conditions, depending on complexity of needs.

**Examples of situation-specific, tailored parts of model**
*(depending on patient/provider/environmental context*

- Patient requires a moderate level of support from care coordinator (e.g. face-to-face appointments with a nurse or equivalent)
- The patient requires regular care appointments, and on demand coordination appointments
- The local healthcare providers and GP have full access to the patient's online records. Letters are sent to local healthcare providers and to the patient to update them.
- Patients may be put in contact with patient support groups where applicable

**Model 8. Patient (adult or child) with an ultra-rare/undiagnosed condition which does not have access to a specialist centre and it is not clear who they need to see + patient/carer cannot coordinate own care**

- The patient will receive care from their local providers only, as there is no specialist service for them to attend.
- A local provider (GP for adults, community paediatrician for children) will act as clinical lead and will oversee the patient's care (e.g. may seek expertise from specialists or refer the patient to quaternary hospitals e.g. geneticists etc.). The clinical lead will work closely with these specialists/quaternary services to manage care (e.g. some options of this include GPs in clinical genetics depts. Or an expansion of the role of geneticists Etc.).
- A formalised care agreement outlining responsibilities, care and team working should be developed (E.g. contracts and access to records).
- The clinical lead will arrange for a local or quaternary provider (E.g. a community matron or nurse at local hospital based on resources) to act as the care coordinator and point of contact for the patient. For undiagnosed patients, this may involve a nurse coordinator who specialises in undiagnosed conditions, depending on complexity of needs.

**Examples of situation-specific, tailored parts of model**

*(depending on patient/provider/environmental context*

- Patient requires a high level of support from care coordinator to coordinate diagnosis, treatments and care (e.g. face-to-face appointments with a SWAN nurse based at a quaternary service, or a GP)
- The patient requires ongoing care appointments, and ongoing coordination appointments, due to their complex care needs and rareness of condition
- The local hospital's infrastructure will be used to share electronic records internally. Letters will be sent to the patients to keep them updated. All healthcare professionals involved in a patient's care will have full access to records (seeing the information that is relevant to them).
- Patients may be put in contact with patient support groups where applicable

**Model 9. Patient (adult or child) with a rare/ultra-rare/undiagnosed condition which does not have access to a specialist centre but it is clear who they need to see + patient/carer has ability and wants to coordinate own care**

- The patient will receive care from their local providers only, as there is no specialist service for them to attend.
- A local provider (GP for adults, community paediatrician for children) will act as clinical lead and will oversee the patient's care. The clinical lead will work with and seek expertise from the relevant disciplines/specialists or refer the patient to quaternary hospitals e.g. geneticists etc. The clinical lead will work closely with these specialists/quaternary services to manage care
- A formalised care agreement outlining responsibilities, care and team working should be developed (E.g. contracts and access to records).
- The healthcare professionals involved in the patient's care will meet (remotely or face-to-face depending on provider locations) to discuss their care.
- The clinical lead will arrange for a local or quaternary provider (E.g. a community matron or nurse at local hospital based on resources) to act as the care coordinator and point of contact for the patient. For undiagnosed patients, this may involve a nurse coordinator who specialises in undiagnosed conditions, depending on complexity of needs.

**Examples of situation-specific, tailored parts of model**

*(depending on patient/provider/environmental context*

- Patient requires a moderate level of support from care coordinator (e.g. face-to-face appointments with a nurse or equivalent)
- The patient requires regular care appointments, and on demand coordination appointments
- The local healthcare providers and GP have full access to the patient's online records. Letters are sent to local healthcare providers and to the patient to update them.
- Mode of appointments, communication and coordination (digital, remote, phone to phone), may vary depending on factors such as situation, resources and patient preferences
- Patients may be put in contact with patient support groups where applicable

**Model 10. Patient (adult or child) with a rare/ultra-rare/undiagnosed condition which does not have access to a specialist centre but it is clear who they need to see + patient/carer cannot coordinate own care**

- The patient will receive care from their local providers only, as there is no specialist service for them to attend.
- A local provider (GP for adults, community paediatrician for children) will act as clinical lead and will oversee the patient's care. The clinical lead will work with andseek expertise from the relevant disciplines/specialists or refer the patient to quaternary hospitals e.g. geneticists etc.. The clinical lead will work closely with these specialists/quaternary services to manage care
- A formalised care agreement outlining responsibilities, care and team working should be developed (E.g. contracts and access to records).
- The healthcare professionals involved in the patient's care will meet (remotely or face-to-face depending on provider locations) to discuss their care.
- The clinical lead will arrange for a local or quaternary provider (E.g. a community matron or nurse at local hospital based on resources) to act as the care coordinator and point of contact for the patient. For undiagnosed patients, this may involve a nurse coordinator who specialises in undiagnosed conditions, depending on complexity of needs.

**Examples of situation-specific, tailored parts of model**

*(depending on patient/provider/environmental context*

- Patient requires a high level of support from care coordinator to coordinate diagnosis, treatments and care (e.g. face-to-face appointments with a SWAN nurse based at a quaternary service, or a GP)
- The patient requires ongoing care appointments, and ongoing coordination appointments, due to their complex care needs and rareness of condition
- The local hospital's infrastructure will be used to share electronic records internally. Letters will be sent to the patients to keep them updated. All healthcare professionals involved in a patient's care will have full access to records (seeing the information that is relevant to them).
- Mode of appointments, communication and coordination (digital, remote, phone to phone), may vary depending on factors such as situation, resources and patient preferences
- Patients may be put in contact with patient support groups where applicable
